# Peripheral blood somatic mosaicism and clonal hematopoiesis across ancestry backgrounds

**DOI:** 10.1101/2025.03.21.25324408

**Authors:** Christelle Colin-Leitzinger, Yi-Han Tang, Mingxiang Teng, Nancy Gillis

## Abstract

Somatic mosaicism (SM), the presence of somatic mutations, is classified as clonal hematopoiesis (CH) when it occurs in hematopoietic cells at an age-related rate. CH is associated with risk for hematologic malignancies and cardiovascular disease, but most studies are predominately based on individuals of European ancestry. Using peripheral blood whole exome sequencing data from 125,748 individuals of diverse genetic ancestries, we cataloged 503,703 SM mutations based on low variant allele frequency distributions and 89,361 CH variants based on age-skewing. We examined CH prevalence across ancestry groups, including commonly recognized pathogenic variants in myeloid (M-CHIP) and lymphoid (L-CHIP) malignancies. CH and M-CHIP variants had the highest prevalence in the European non-Finnish ancestry group, and males trended toward more M-CHIP variants. Ancestry differences in CH included more mutations in *NF1* in African/African American, *TP53* in European, and *CUX1* in Asian and Latino/Admixed American ancestry groups. Linking the identified CH variants to a cancer database, CH was detected in 14% (55,190/391,102) of patient tumors. Prevalence of CH variants in some solid tumors ranged from 25% - 40%. M-CHIP variants in solid tumors were associated with younger age (61 vs 63, p <0.001), while M-CHIP in hematologic malignancies were linked to older age (60 vs 50, p <0.001), suggesting differences in disease biology. This study provides a catalog of SM, CH, and CHIP variants across diverse ancestry groups, highlighting differences that are important to inform clinical care, drug discovery, and study design to maximize generalizability across individuals.

**KEY POINTS:**

- Establish a large-scale catalog of somatic mosaicism and clonal hematopoiesis mutations across genetic ancestry groups.
- Mutation occurrences within clonal hematopoiesis genes vary across ancestry groups, with age, and across tumor types.

## INTRODUCTION

Somatic mosaicism (SM), the accumulation of somatic mutations, begins during embryonic development and occurs in tissues and cells throughout the body. SM is particularly common in tissues with high rates of cell turnover, such as the skin, liver, and hematopoietic cells.^1^ Age-associated SM in hematopoietic cells has been referred to as clonal hematopoiesis (CH).^2,3^ Although most SM and CH alterations will not cause disease, some can cause cancer or increase risk for cardiovascular disease, macular degeneration, or neurological conditions.^4–8^ In fact, due to broad implications and poorly understood effects of SM, the National Institutes of Health launched the Somatic Mosaicism Across Human Tissues (SMaHT) initiative to catalogue these mutations and investigate their impact on human health and disease.^9^

In peripheral blood, SM is most well-studied in the context of CH. CH becomes increasingly common as individuals age, and is associated with over ten-fold increased risk for hematologic malignancies, twice the likelihood of cardiovascular disease, and approximately 40% increased risk of all-cause mortality.^10–12^ Risk for these adverse outcomes has been linked to SM primarily in genes recurrently mutated in hematologic malignancies (e.g., *DNMT3A*, *TET2*, *ASXL1*, *JAK2*, etc.), referred to as clonal hematopoiesis of indeterminate potential (CHIP) when the variant allele frequency (VAF) is at least two percent.^3,13^ As such, the extensive literature on SM in peripheral blood largely focuses on only these genes. Two sets of CHIP candidate genes exist, representing different disease risks: myeloid malignancy-related (M-CHIP) and lymphoid malignancy-related (L-CHIP).^14^ Limiting research to these candidate gene lists has benefits, such as reducing multiple testing and increasing likelihood of statistically significant findings; however, broad use of this approach provides limitations to our comprehensive understanding of SM in peripheral blood. Another notable limitation of this approach is that the CHIP genes commonly studied are most commonly mutated in White or European individuals, the populations in which they were discovered, and less common in racial and ethnic minority groups.^11,15^ Therefore, SM in peripheral blood of diverse racial and ethnic populations is even less understood.

Large-scale sequencing studies provide opportunities to identify SM variants across populations. Here, we use the Genome Aggregation Database (gnomAD) to catalogue SM and CH variants in the peripheral blood from individuals with diverse ancestry backgrounds. The gnomAD population is one of the largest collections of harmonized whole exome sequencing (WES) data aggregated from over 60 studies, which has been reprocessed through a unified pipeline.^16^ The repository contains variant data from 125,748 exomes and genetic ancestry representation with allele frequency information from five continental groups (African/African American, East Asian, European, Latino, and South Asian), two demographically distinct groups (Ashkenazi Jewish and Finnish), and uncategorized (Other) samples. In this study, we analyzed SM and CH using peripheral blood WES data from the aggregated gnomAD population, examining differences in prevalence and mutated genes across genetic ancestry groups. We contextualized our findings within existing literature, focusing on commonly studied CHIP genes (M-CHIP and L-CHIP).^3,15^ Additionally, we interrogated the gnomAD results in the context of cancer by linking findings to the Catalogue Of Somatic Mutations In Cancer (COSMIC) database.^17^ This study provides a novel catalogue of SM in peripheral blood across diverse ancestry groups, serving as a first step toward understanding the clinical impact and disparities associated with these alterations.

## METHODS

### Data sources

Aggregate WES data derived from peripheral blood was downloaded from gnomAD (v2.1.1), lifted over from the GRCh37 to the GRCh38 reference sequence (https://gnomad.broadinstitute.org/downloads).^16^ Extracted data elements critical for this study include age, allele balance (AB), allele count (AC), mutation allele depth (AD or depth), and population allele frequency (PAF) (defined in **Table 1**). Ancestry groups as defined by gnomAD with African/African American, Ashkenazi Jewish, East Asian, European (Finnish), European (non-Finnish), Latino/Admixed American, and South Asian ancestry were included in this study (**Table S1**). The non-cancer population subset, only annotated in gnomAD v2.1.1, was used for comparing variant PAFs across ancestry groups.

**Table 1.**
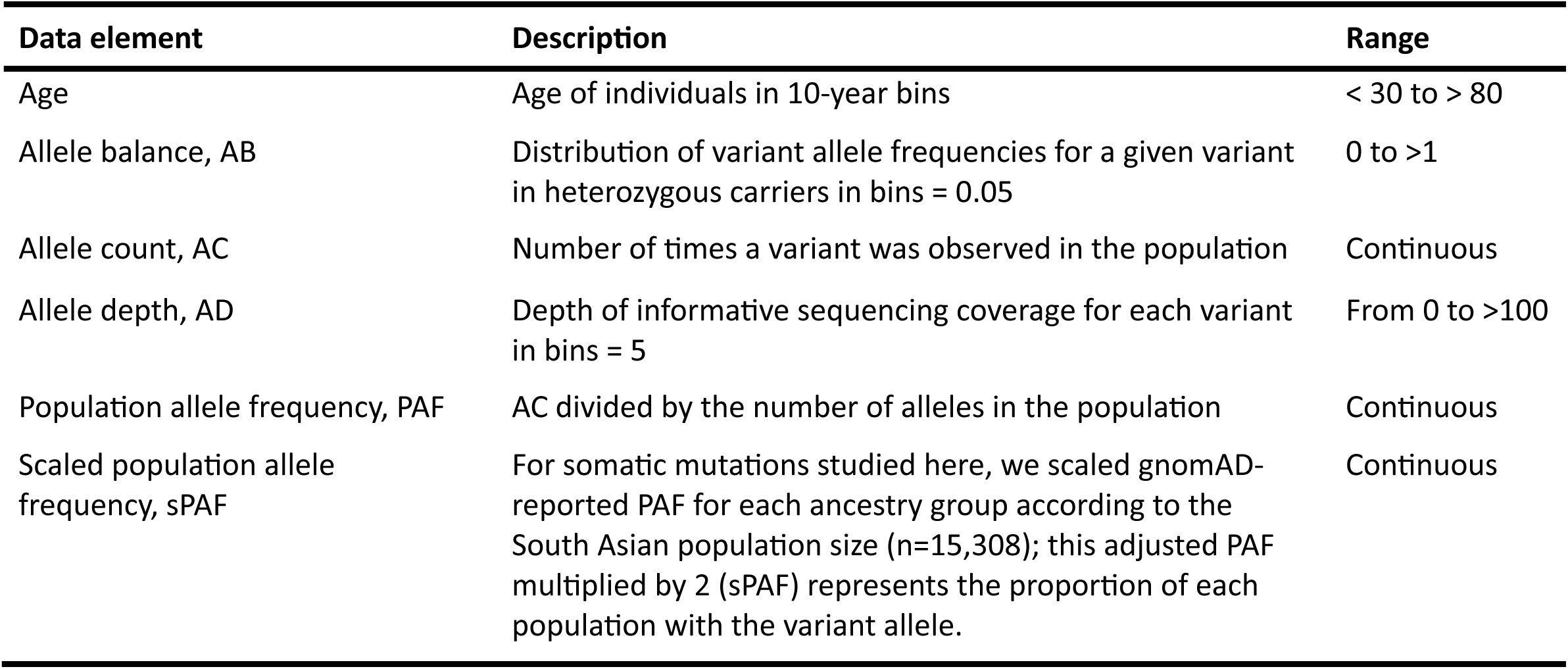
Data elements.

To explore SM and CH variants in the context of potential cancer pathogenicity, data from gnomAD was linked to Catalogue of Somatic Mutations in Cancer (COSMIC, v95, https://cancer.sanger.ac.uk/cosmic/download) using variant chromosome number, position, reference, and alternate alleles.^17^ Additional data elements extracted from COSMIC were mutation description, mutation somatic status, FATHMM prediction of pathogenicity,^18^ age at sample collection, cancer primary site, and histology. SM and CH variants identified in gnomAD with an occurrence > 1 time in COSMIC were included in the COSMIC-related analyses.

### Identification of somatic mosaicism and clonal hematopoiesis variants

Variants in the gnomAD database were considered if AB information was available from more than five individuals (n=3,934,541 variants; **Figure 1A**). We applied stringent quality control filters to eliminate artifacts and low-quality variants. First, variants detected exclusively within the first AB bin (0.00 - 0.05) were considered background noise or sequencing artifacts and removed from further analysis. Variants with a median depth < 10 were removed due to poor coverage and inability to accurately detect SM or CH, which are characterized by low VAFs. Variants located in difficult-to-sequence regions, as annotated by the UCSC table browser,^19^ were also removed, including low complexity regions (WindowMasker + SDust^20^), segmental duplications, centromeres, and variants in the blacklisted regions developed by ENCODE.^21^ We further restricted analysis to include only protein coding genes, as defined by GENCODE v42 annotation.^22^

**Figure 1.**
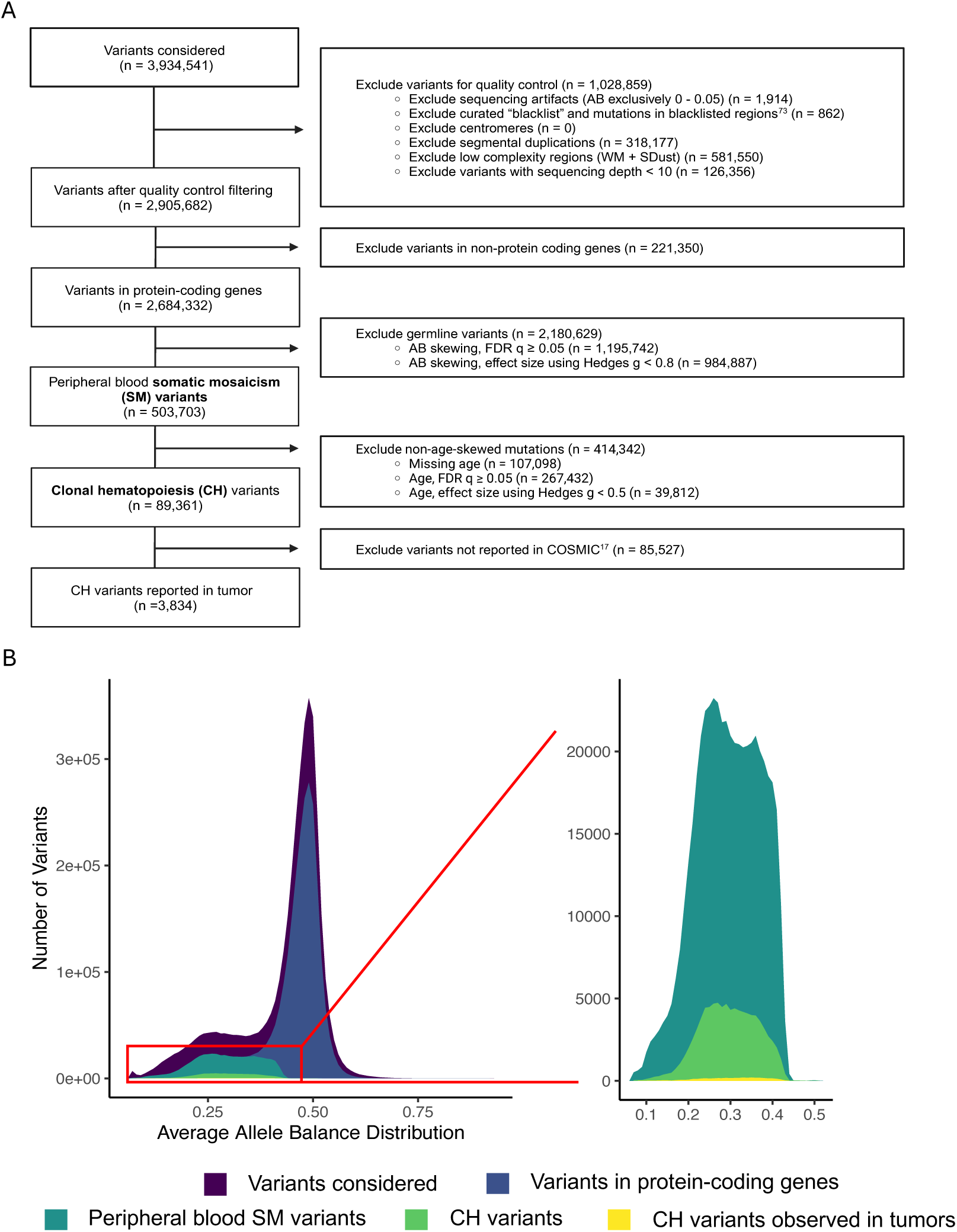
Identification of somatic mosaicism (SM) and clonal hematopoiesis (CH) variants using the gnomAD population. **(A)** Consort diagram representing the successive filters and methods applied to identify variants of interest. Variants with AB information available from more than five individuals were considered for inclusion. **(B)** Distribution of allele balance for the consecutive variant subsets.

SM variants were selected based on AB skewing, a function of VAF. Specifically, variants in the gnomAD population with consistently low VAF (e.g., <25%) were assumed to be somatic mutations, as opposed to germline mutations which have VAFs centered around 50%. To identify AB skewing, a one-sided Mann-Whitney test was conducted to compare each variant’s AB distribution to the AB distribution of a reference mutation (defined as AB from a common germline variant in *DNMT3A*, rs2276599) (**Figure 2A**). Statistical significance was adjusted for multiple testing using the Benjamini-Hochberg’s FDR method.^23^ Hedges g statistic was used to quantify the extent of skewing (i.e., the effect size).^24^ SM variants were selected with an AB skewed toward lower values with an FDR q < 0.05 and Hedges g ≥ 0.8 (i.e., large effect size).

**Figure 2.**
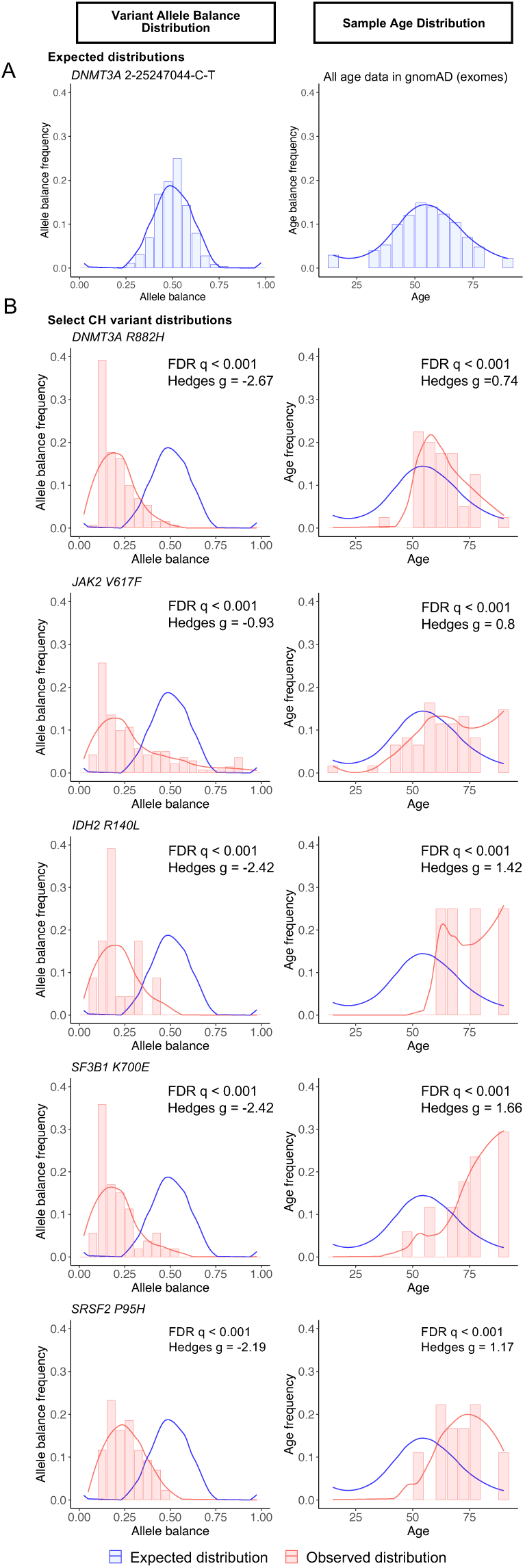
Somatic mosaicism (SM) and clonal hematopoiesis (CH) variant identification. **(A)** Expected distributions of allele balance (AB, left) and age (right). AB reference was based off the germline mutation *DNMT3A* chr2-25247044-C-T (rs2276599); age reference was estimated based on ages of the aggregate gnomAD population. **(B)** Examples of CH variants identified due to skewing of AB (left) toward lower values and age (right) toward higher values. Blue curves show the expected distributions, and red curves show the observed variant data.

CH variants were selected from the SM variants with the added characteristic that their prevalence was skewed toward older individuals when compared to the aggregate gnomAD population, which has a normal-like age distribution. The same statistical methods were used to classify age skewing as described for AB skewing (i.e., Mann-Whitney tests with FDR correction). To identify CH variants, SM variants were tested for age distribution skewing toward higher ages (FDR q < 0.05 and Hedges g ≥ 0.5) compared to the aggregate gnomAD population (**Figure 2A**). CH variants were annotated using ANNOVAR,^25^ which included extraction of the combined annotation-dependent depletion (CADD) score,^26^ and linked to COSMIC. PAFs of CH variants were also extracted from the dbGaP population (dbSNP build 155).^27,28^ Presence of CH variants in M-CHIP and L-CHIP gene regions (**Table S2**) were identified based on the gene, amino acid change, function, and exonic function annotation.^3,14,15^

The PAF information from gnomAD was used to evaluate differences in SM and CH variant prevalence across ancestry groups. Overall PAFs were compared using the Kruskal-Wallis test. To mitigate differences in ancestry group sizes, PAFs were scaled to the size of the South Asian ancestry group (the middle ancestry group size). Then, to estimate person-level prevalence from allele-count frequencies (PAF), gnomAD-reported PAFs were multiplied by two; this was defined as the scaled PAF (sPAF) (**Table 1**). Upset plots were used to show gene and loci overlap across ancestry groups. Wilcoxon rank-sum tests were used to compare sPAFs stratified by sex and age in the COSMIC data. To guide future SM and CH studies that use population-based data (e.g., gnomAD) as a filter for removing germline mutations from data, we also calculated a PAF cutoff to distinguish likely germline from likely CH variants. The cutoff was chosen so that it has an equal number of standard deviations from the mean of the whole gnomAD PAF distribution and the mean of the CH-only PAF distribution. All statistical analyses were done using R version 4.3.0.

## RESULTS

### Selection of variants for inclusion

This study analyzed aggregate data from 125,748 human whole exomes, which captured 3,934,541 variants with AB information available in more than five individuals (**Figure 1A**). For these variants, AB distribution was bimodal, with most variants having an AB centered around 0.5 and a minority of variants with an AB centered around 0.25 (**Figure 1B**). Quality control filtering removed almost one-third of the variants (n=1,028,859, **Figure S1**). Most of the variants removed were in low complexity regions of the genome or segmental duplications. Of the remaining variants, 2,684,332 (92.4%) were in protein-coding genes and included in downstream analysis (**Figure 1A**). The AB distribution of the variants in protein-coding genes was comparable to the initial dataset, with a large peak near 0.50 and a smaller group with an AB close to 0.25 (**Figure 1B**).

### Somatic mosaicism and clonal hematopoiesis variants

A total of 503,703 peripheral blood variants were classified as SM based on AB skewing (**Figure 1, Table S3**). Of the SM variants, 107,098 (21.3%) were derived from studies that did not have age data in gnomAD, so could not be assessed as CH variants. Of the remaining SM variants, 89,361 (of 396,605 or 22.5%) also had age-related skewing and were classified as CH (**Figure 1, Table S3**). Of the variants reported in gnomAD, 12.8% and 2.3% were classified as SM and CH, respectively. SM variants showed AB skewing, while CH variants had AB and age skewing (**Figure 2**). We statistically modeled PAF distributions to determine population frequency estimates for germline versus somatic mutations, and a PAF cutoff of 2.4 x 10^-5^ was estimated for defining likely somatic mutations in gnomAD.

Overall, the types of variants identified as SM and CH were similar, with most being nonsynonymous single nucleotide variants (SNVs), followed by intronic, and synonymous SNVs (**Figure 3A**). The intronic variants are expected in regions closely flanking the targeted exons. SNVs were enriched in SM and CH compared to the entire gnomAD dataset (**Figure S2**). Additionally, cytosine-to-thymine (C→T) transitions were most common for both SM and CH variants (58.7% and 60.9%, respectively; **Figure 3B**), as expected in CH and aging.^11,29^ Of the SM and CH variants identified, only 6.2% and 5.6%, respectively, are predicted to be somatic according to Ensembl variant effect predictor annotation in gnomAD;^30^ however, most of the SM and CH variants (64% and 65%, respectively) have a CADD score classified as likely pathogenic (i.e., ≥ 20), indicating they are predicted to be among the 1% most deleterious mutations (**Table S4**).^26^ The prevalence of SM variants across all genes exhibited a gradual increase until approximately 70 years of age, after which it showed a slight decline. In contrast, CH variant prevalence steadily rose with age, with a notable change in slope occurring around 60 years old (**Figure 3C**).

**Figure 3.**
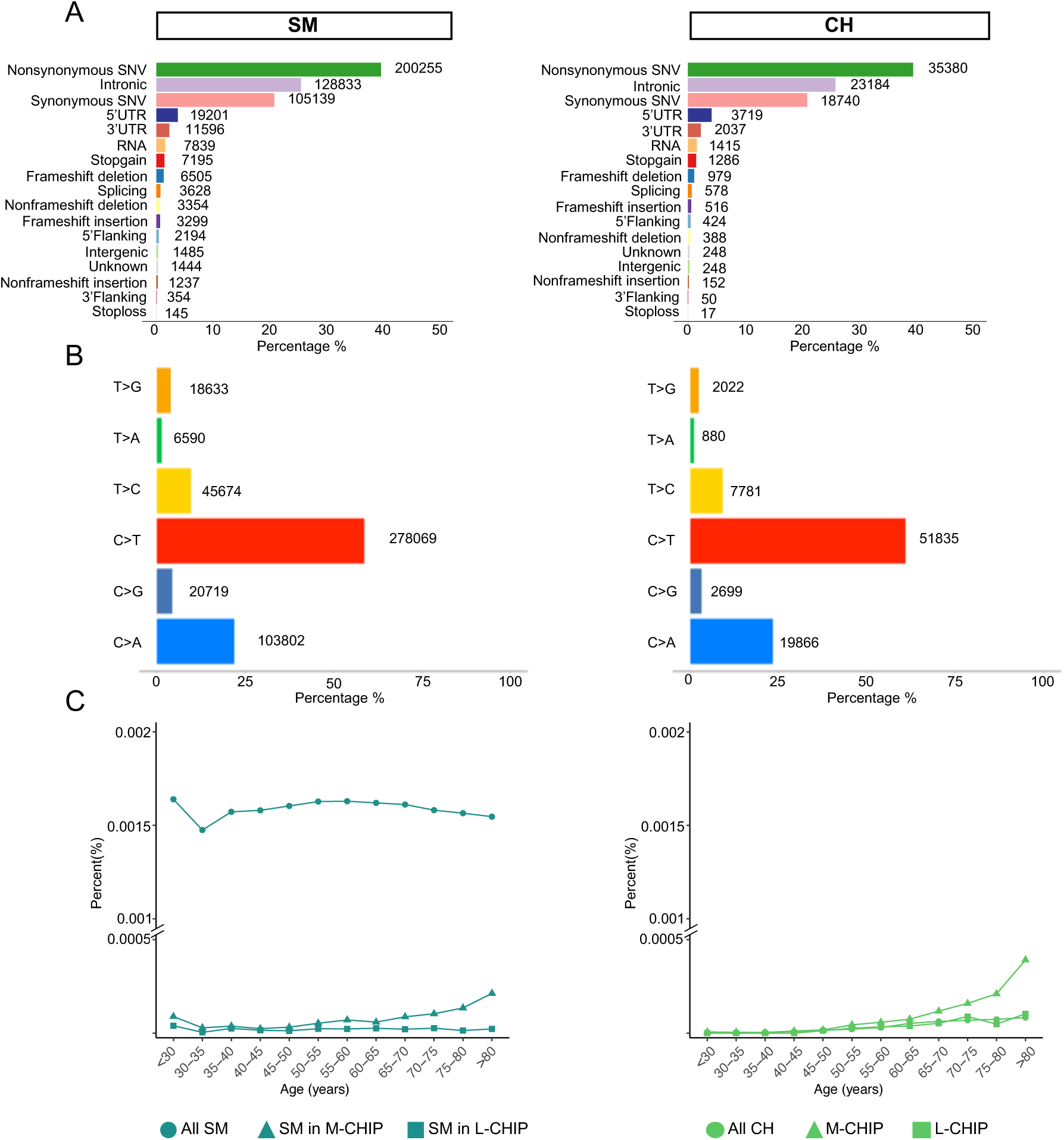
Overall characteristics of the identified somatic mosaicism (SM) and clonal hematopoiesis (CH) variants. Figure shows the **(A)** functional characterization, **(B)** frequencies of nucleotide transitions, and **(C)** age distributions of the SM and CH variants. Age distributions were plotted based on the binned age information in gnomAD. Percent on the y-axis indicates the percent of loci tested that had a SM or CH mutation for a given age group.

To assess the frequency of SM and CH across genes, we summed AC to determine the total occurrences of variants within each gene, enabling comparative analyses across genes (**Table S5**). Among M-CHIP genes, the genes with the most SM variants were *DNMT3A*, *CUX1*, *SF3B1*, *NF1*, and *TP53*; the genes with the most CH variants were *DNMT3A*, *JAK2*, *MFSD11*, *SF3B1*, and *PDS5B* (**Table S4**). Among L-CHIP genes, the genes with the most SM variants were *TSC2*, *SMARCA4*, and *KMT2D*; the genes with the most CH variants were *SMARCA4*, *KMT2D*, *NOTCH1*, and *POT1* (**Table S4**). Results were consistent when examined in the non-cancer subset of the gnomAD population.

#### Somatic mosaicism and clonal hematopoiesis variants across sex, ancestry, and age groups

Ancestry groups in this study were based on availability in gnomAD and included European non-Finnish (n=56,885, 45.2%), Latino or Admixed American (n=17,296, 13.8%), South Asian (n=15,308, 12.2%), European Finnish (n=10,824, 8.6%), East Asian (n=9,197, 7.3%), African or African American (n=8,128, 6.5%), and Ashkenazi Jewish (n=5,040, 4%) (**Table S1**). There were slightly more males (n=67,961, 54%) than females (n=57,787, 46%) included.

The genes with SM and CH variants largely overlapped across ancestry groups; however, SM and CH-mutated individual loci were more variable across ancestry groups (**Figure 4**). The European non-Finnish ancestry group had the largest number of unique SM and CH loci, followed by South Asian for SM and Latino/Admixed American for CH (**Figure 4B**). The median sPAF of SM variants was highest in European Finnish and Ashkenazi Jewish ancestry groups; the highest sPAF of CH variants was in European non-Finnish ancestry group followed by Ashkenazi Jewish (**Figure 4C and Table S6**). The high sPAF value indicates that the European non-Finnish ancestry group has more CH variants that are common among individuals than other ancestry groups, where individual variants occur in fewer individuals. Although SM variants were more common in males than females (p<0.001), the prevalence of CH variants was similar in both sexes (p=0.2) (**Table S6**).

**Figure 4.**
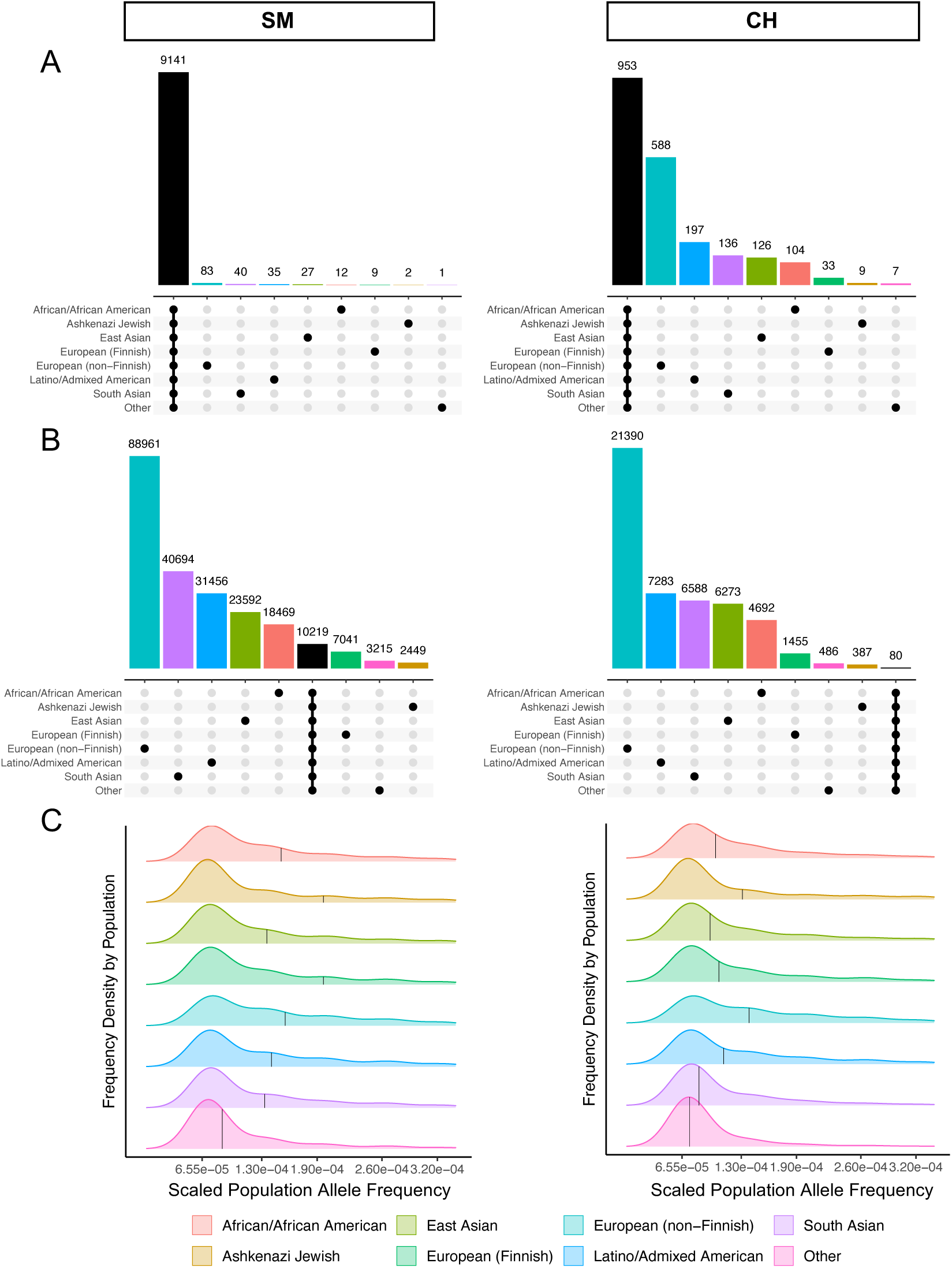
Somatic mosaicism (SM) and clonal hematopoiesis (CH) variants across ancestry backgrounds. Upset plots depicting the **(A)** number of genes and **(B)** individual loci with SM and CH shared across ancestry groups. Black bars show the overlap between ancestry groups. **(C)** Density plot of the scaled population allele frequencies (sPAF) of SM and CH variants in non-cancer individuals by ancestry group. Black vertical lines show the median sPAF in each ancestry group.

### Somatic mosaicism and clonal hematopoiesis in known pathogenic regions

To understand ancestry differences in mutations associated with hematologic malignancies, we focused on curated gene regions. The sPAF in M-CHIP curated regions was highest for the European non-Finnish ancestry group, and males trended toward more M-CHIP variants than females (**Figure 5**, **Table S7**). *DNMT3A* was consistently among the highest SM-mutated M-CHIP genes, but beyond that, differences occurred across ancestry groups (**Figure 6, Table S8**). For example, *NF1* had the most SM variants overall in the African/African American ancestry group and was second-most SM-mutated gene in the East Asian ancestry group, while *TP53* was the second-most SM-mutated gene in the European non-Finnish ancestry group, and *CUX1* was the most SM-mutated gene in the South Asian ancestry group and second-most SM-mutated gene in the Latino/Admixed American ancestry group. The M-CHIP genes with CH were similar across ancestry groups, with *DNMT3A* and *JAK2* being most common. *SF3B1* showed a trend of a higher overall frequency of CH variants in the Latino/Admixed American ancestry group compared to other ancestry groups.

**Figure 5.**
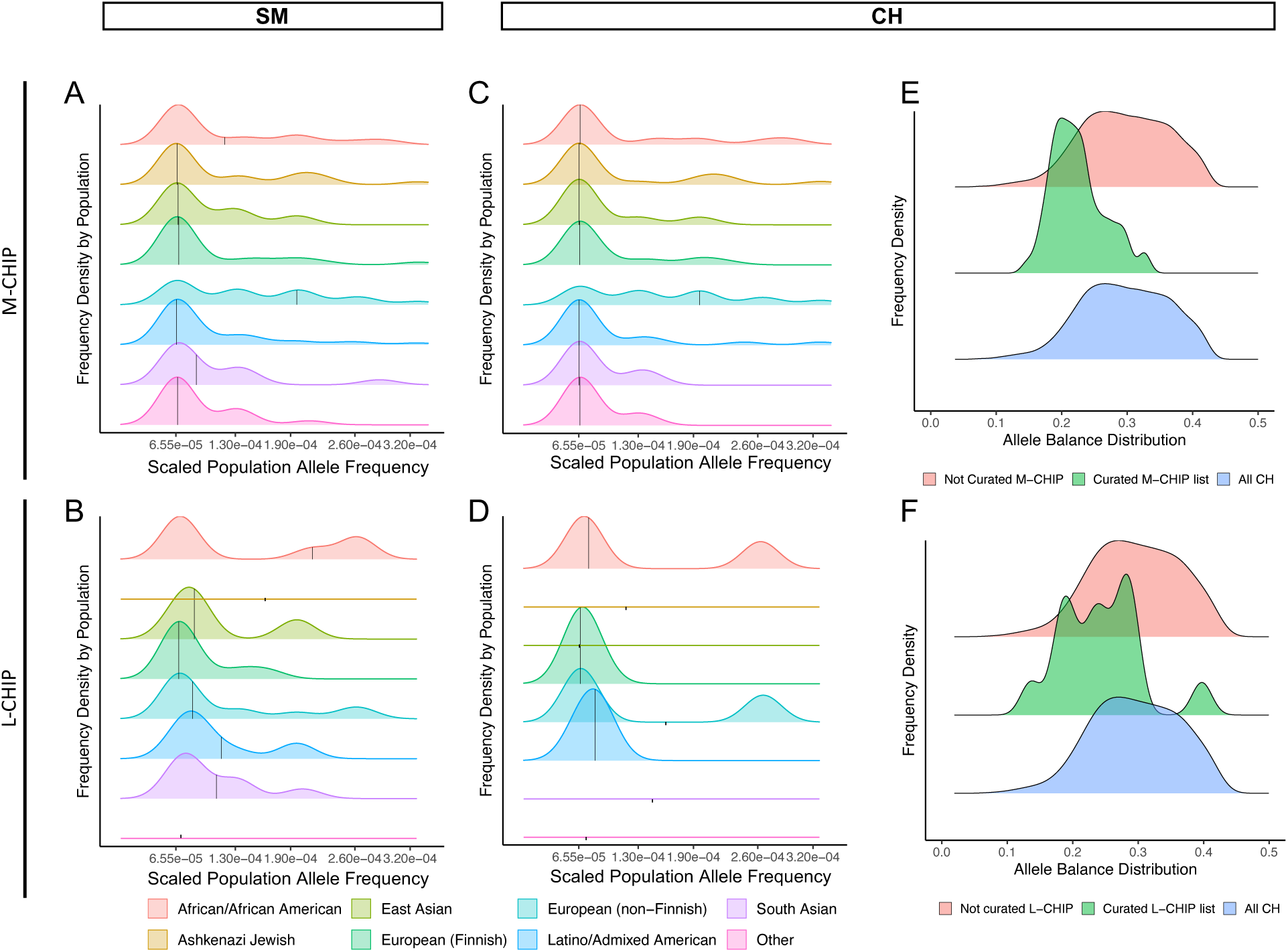
Somatic mosaicism (SM) and clonal hematopoiesis (CH) in curated CHIP regions. Density plots show the scaled population allele frequency (sPAF) distributions of **(A, B)** SM and **(C, D)** CH variants in myeloid (M-CHIP) and lymphoid (L-CHIP) curated regions across ancestry groups. Black vertical lines show the median sPAF in each ancestry group. Allele balance distributions **(E, F)** represent the allele frequencies of CH stratified by occurrence in curated gene regions.

**Figure 6.**
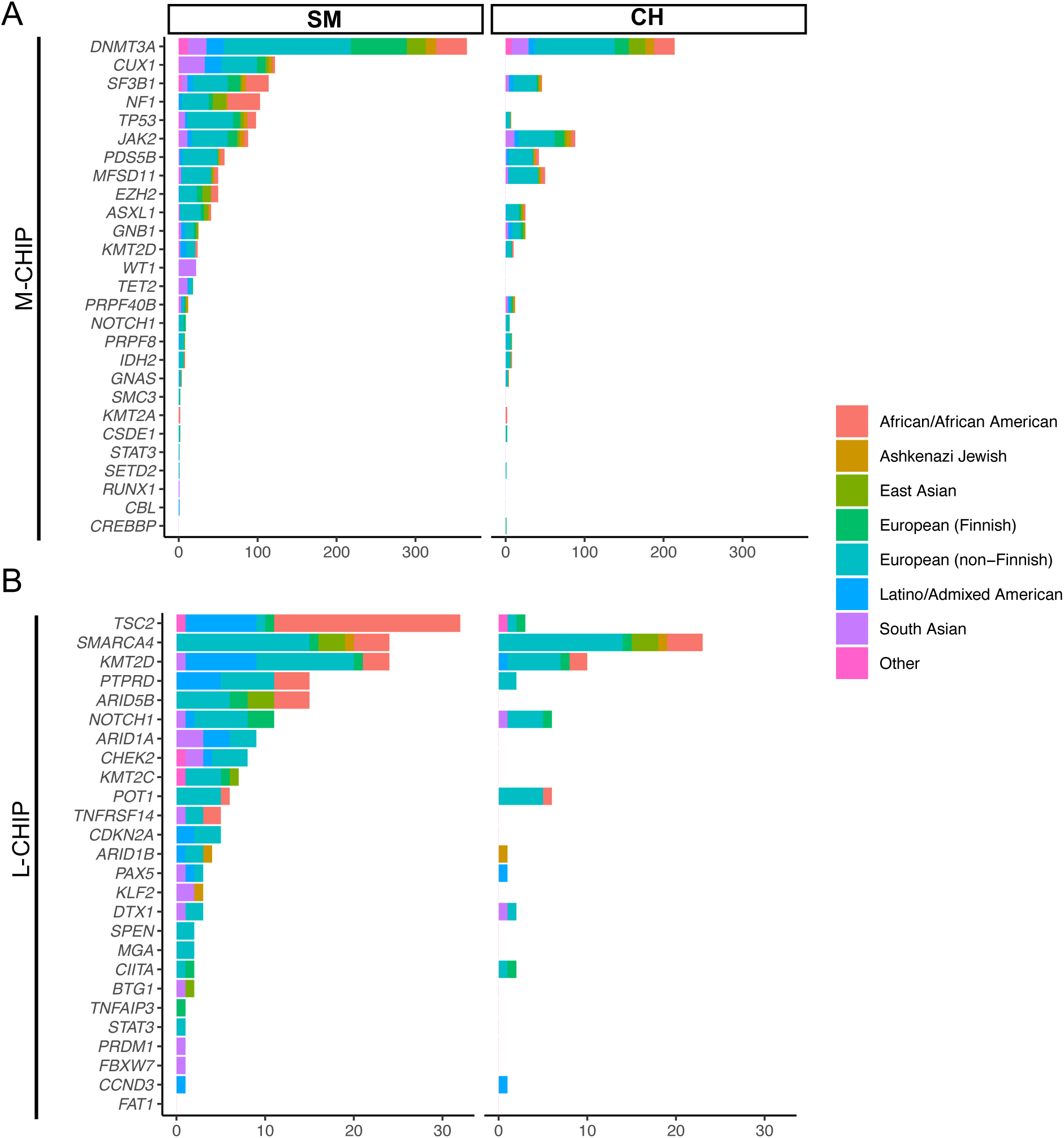
Prevalence of somaTc mosaicism (SM) and clonal hematopoiesis (CH) variants within the curated gene regions. **(A)** Data based on myeloid (M-CHIP) and **(B)** lymphoid (L-CHIP) gene regions. Prevalence is estimated based on the cumulative variant allele count within each gene.

SM and CH in L-CHIP regions are less common than in M-CHIP regions, which limits power for understanding differences across ancestry groups, but differences were noted. The highest sPAF of SM variants in L-CHIP regions was in the African/African American followed by the Ashkenazi Jewish and Latino/Admixed American ancestry groups (**Figure 5**, **Table S7**). There were no differences in L-CHIP by sex. The L-CHIP genes with the most SM variants overall were *TSC2*, *SMARCA4*, and *KMT2D*. Differences by ancestry included: *SMARCA4* and *KMT2D* had the most SM in the European non-Finnish ancestry group, *TCS2* in the African/African American ancestry group, and *KMT2D* and *TCS2* in the Latino/Admixed American ancestry group (**Figure 6**, **Table S9**). CH in L-CHIP regions was most common in *SMARCA4* overall, driven by its prevalence in the European non-Finnish, African/African American, and East Asian ancestry groups.

The average AB distributions for M-CHIP and L-CHIP variants were lower than the overall group of CH variants (**Figure 5**), indicating consistently lower VAFs for known regions. Looking at curated CHIP mutations, most variants (e.g., JAK*2 V617F*, *DNMT3A R882\**, *SRSF2 P95\**, *GNB1 K57E* and SF*3B1 K700E*) have the highest prevalence in the European non-Finnish ancestry group (**Figure S3**). Interestingly, for the most common CHIP mutation known, *DNMT3A R882H*, the East Asian ancestry group has the second highest prevalence, followed by African/African American. *JAK2 V16F* is most common in the European Finnish, followed by South Asian ancestry groups. *ASXL1 Y591X* was only detected in the European non-Finnish ancestry groups.

### CH variants in tumors and curated CHIP gene lists

To explore the presence and potential impact of CH mutations in cancer, we examined the CH variants identified from gnomAD in COSMIC. Of the CH variants identified in gnomAD, 4.3% (3,834/89,361) were present in 14% of patient tumors (55,190/391,102 unique patients) (**Figure 1**). Patients with CH variants detected in their tumors were older than those without CH (median age 61 vs 59; p <0.001; **Figure S4, Table S10**). This trend persisted when stratifying patients by hematologic (60 vs 51) and non-hematologic (62 vs 60) cancers (p <0.001). CH variants were detected across cancer types, with the vast majority occurring in hematopoietic cancers (85%) and more commonly in those with hematopoietic (97%) than lymphoid (1.8%) neoplasm histology (**Table 2**). Within hematologic cancers, most patients (87%) carried gnomAD-curated CH mutations (**Figure 7, Table S11**). Among solid cancers, CH variants were most common in large intestine (41%), endometrium (38%), stomach (35%), skin (32%), pancreas (28%), and thyroid (26%) tumors (**Figure 7, Table S11**). Like gnomAD, the CH variants observed in COSMIC were mostly missense or nonsynonymous (95%); however, they were mostly classified as pathogenic (91%) and reported or confirmed as somatic variants (99%) in COSMIC (**Table 2**).

**Figure 7.**
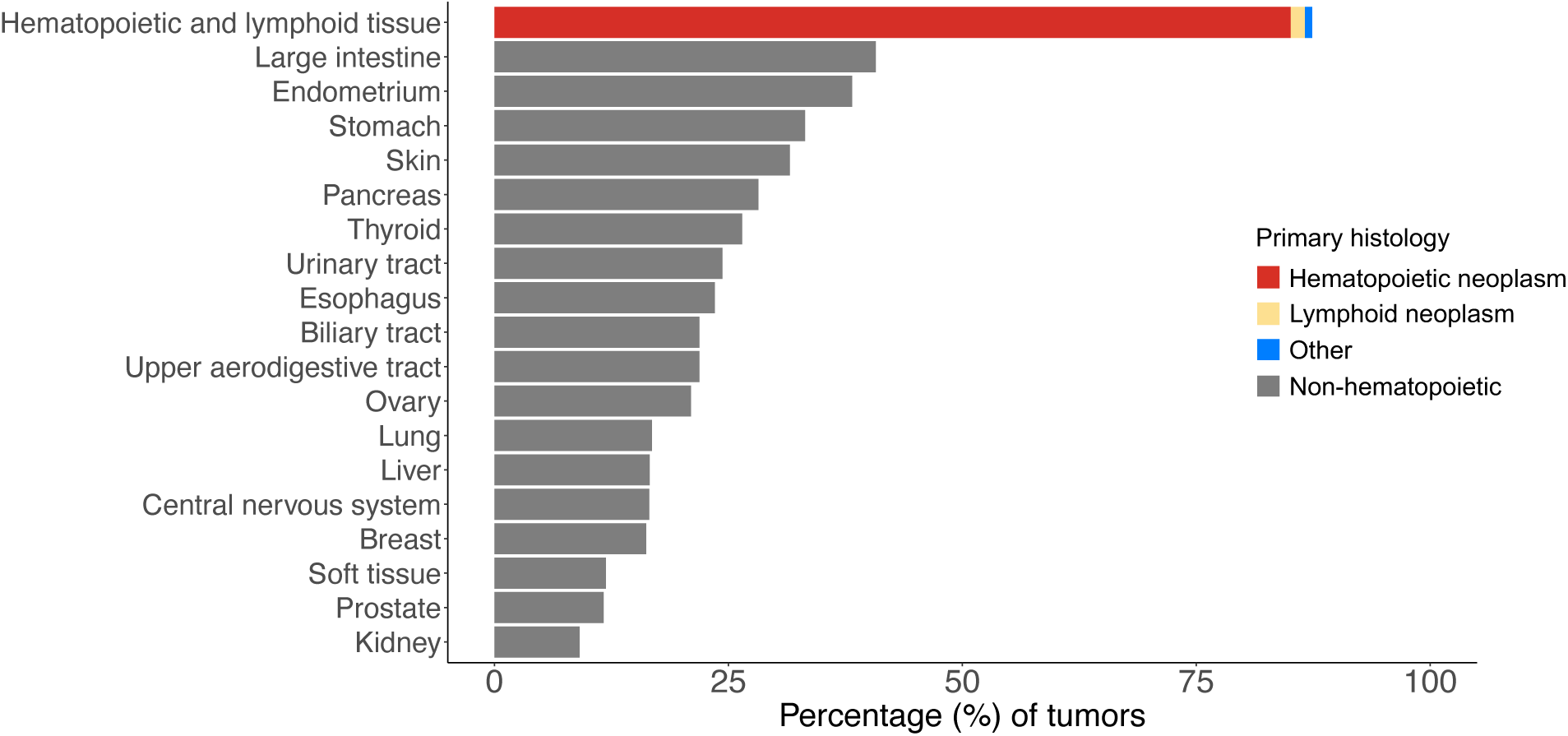
Prevalence of clonal hematopoiesis (CH) across tumor types in COSMIC. All tumor types with more than 500 samples in COSMIC are shown.

**Table 2.** Characteristics of clonal hematopoiesis (CH) in patient tumor samples. Cancer types with more than 500 samples are included. Individuals are considered as CH carriers if at least one CH mutation is detected.

| COSMIC samples, N =55,190 |  |
| --- | --- |
| **Primary site, n (%)** |  |
| Hematopoietic and lymphoid tissue | 46,744 (85%) |
| Hematopoietic neoplasm | 45,500 (97%) |
| Lymphoid neoplasm | 826 (1.8%) |
| Other | 418 (0.9%) |
| Large intestine | 1,802 (3.3%) |
| Skin | 633 (1.1%) |
| Lung | 604 (1.1%) |
| Breast | 582 (1.1%) |
| Pancreas | 538 (1.0%) |
| Stomach | 505 (0.9%) |
| Central nervous system | 391 (0.7%) |
| Liver | 347 (0.6%) |
| Upper aerodigestive tract | 345 (0.6%) |
| Endometrium | 336 (0.6%) |
| Esophagus | 319 (0.6%) |
| Prostate | 294 (0.5%) |
| Urinary tract | 244 (0.4%) |
| Ovary | 243 (0.4%) |
| Thyroid | 236 (0.4%) |
| Biliary tract | 186 (0.3%) |
| Bone | 160 (0.3%) |
| Kidney | 149 (0.3%) |
| COSMIC mutations, N =61,648 |  |
| **Mutation description, n (%)** |  |
| Substitution - Missense | 55,476 (95%) |
| Substitution - Coding silent | 2,458 (4.2%) |
| Substitution - Nonsense | 473 (0.8%) |
| Nonstop extension | 6 (<0.1%) |
| Unknown | 3,235 |
| **FATHMM prediction, n (%)** |  |
| Pathogenic | 55,635 (91%) |
| Neutral | 5,270 (8.7%) |
| Unknown | 743 |
| **Mutation somatic status, n (%)** |  |
| Reported or confirmed somatic | 61,264 (99%) |
| Variant of unknown origin | 384 (0.6%) |

Across COSMIC tumors, patients with an M-CHIP variant were younger than patients without an M-CHIP variant (median age 61 vs 62, p < 0.001, **Table S12**). This difference persisted when considering only non-hematologic tumors (median age 61 vs 63, p <0.001). On the contrary, in hematologic cancers, patients with a variant in an M-CHIP region were significantly older than those without M-CHIP (median age 60 vs 50, p <0.001). Patients in COSMIC with a variant in an L-CHIP region were younger in all cancer types (median 50 vs 61 years) and hematologic cancers (median 19 vs 60 years), but the sample sizes were too small to make meaningful interpretations.

## Discussion

Although designed with the intent of driving germline genetic association studies, computational advances and the large-scale nature of population databases allow for investigation of rare or less common mutations. The allele balance distribution of the data in gnomAD (Figure 1B) closely mirrors that seen in seminal CH studies,^10^ indicating utility of this population database for studying acquired mutations, such as SM and CH. Using allele imbalance, or low VAFs, we identified 503,703 SM variants and, by incorporating older age skewing, 89,361 CH variants in 125,748 ancestrally diverse individuals. The characteristics of the SM and CH variants identified were as expected based on existing knowledge – overrepresentation of SNVs and cytosine-to-thymine (C→T) transitions, which are most common in CH and drive the somatic mutational signature of aging.^11,29^ This further substantiated our approach and the association between somatic mutation acquisition and aging.

After adjusting for differences in the gnomAD population sizes, SM allele frequencies were highest in the European Finnish and Ashkenazi Jewish ancestry groups, while CH allele frequencies were highest in the European non-Finnish group. This finding indicates that there are more SM variants common among individuals with European Finnish and Ashkenazi Jewish ancestry, the most genetically homogenous ancestry groups included, while there are more CH variants that are common in individuals with European non-Finnish ancestry. The implication of this result is that larger sample sizes may be needed to quantify risk of CH variants in ancestry groups other than European non-Finnish since the likelihood of having multiple individuals with the same variants is much lower. Furthermore, SM and CH variants in the known pathogenic regions had lower VAFs than variants not contained within the curated gene lists. This finding is in line with expectations, since mutations with greater allelic skewing are more likely to be discovered in smaller, less diverse studies due to larger separation from the median. Overcoming the challenge of uncommon variants is further exacerbated by the low representation of diverse individuals in genetic studies and clinical trials.^31,32^

Using a curated set of CH regions related to myeloid malignancies (M-CHIP), SM and CH were most frequent in *DNMT3A* in the overall population. All SM variants in *JAK2* had age skewing and were classified as CH, making it the second most mutated CH gene. Interestingly, two of the genes with the highest prevalence of CH variants (*MFSD11* and *PDS5B*) are not commonly annotated in CHIP studies but do play a pathogenic role in hematopoietic cancers.^33–35^ Knowledge of these mutations and population frequencies can inform prioritization for inclusion in future studies investigating clinical impact and disease biology. Using the same approach with a curated set of CH genes related to lymphoid malignancies (L-CHIP), CH prevalence was low, limiting our ability to identify meaningful differences. L-CHIP SM was most frequent in the tumor suppressor gene *TSC2*, while CH was most common in *SMARCA4* and *KMT2D*.

SM and CH characteristics were different across ancestry groups. Notable differences include high rates of *CUX1* SM variants in Latino/Admixed American and South Asian individuals and *NF1* SM variants in African/African American and East Asian individuals, both genes of which are associated with pathogenesis of hematopoietic cancers.^36–38^ Considering CH, the Latino/Admixed American ancestry group had a higher prevalence of CH variants in *GNB1* compared to other ancestry groups. *GNB1* mutations are postulated to drive initiation of CH and clinical progression of disease.^39^ CH variants in *KMT2A* (or *MLL*) were only detected in the African/African American ancestry group. These differences highlight potential for unique biology and pathogenesis for myeloid diseases across ancestry groups.

We previously showed that blood-derived CH mutations are frequently detected in clinical sequencing of patient tumors^40^ and can impact the tumor-immune microenvironment to affect patient outcomes.^41–47^ CH has also been associated with an increased risk of some solid cancers.^48^ When cross-referencing the CH variants identified in gnomAD with tumor sequencing data in COSMIC,^17^ 14% of the tumors sequenced had a detectable CH mutation and the prevalence was significantly higher in some cancer types. For example, approximately 85% of hematologic cancers had a CH mutation, and prevalence ranged from approximately 25% to over 40% for some solid tumors, including large intestine, endometrium, stomach, skin, pancreas, and thyroid. These rates may underrepresent the true prevalences of CH in tumors, since not all CH genes or variants were identified in gnomAD due to missing age data needed to identify age skewing (i.e., 21% of the SM variants are missing age data in gnomAD). Also, without paired blood sequencing we also cannot confirm that the mutations were in fact blood-derived CH rather than tumor-derived mutations. Nonetheless, these results provide reasonable estimates of the relative rates and impact of CH across cancer types. Further work deciphering the impact of CH within the tumor microenvironment is important to understand disease biology and inform cancer prevention and personalized treatment strategies.

Interesting patterns in the ages of patients with CHIP mutations in their tumors were identified. Overall, patients with M-CHIP and L-CHIP in their tumors were chronologically younger than those without M-CHIP or L-CHIP. This may suggest that individuals with CHIP are biologically more aged than those without CHIP, leading to earlier risk for age-related disease such as cancer. In fact, CH has been linked to epigenetic age acceleration in multiple non-cancer cohort studies.^49–51^ On the contrary, when considering only patients with hematologic malignancies, those with M-CHIP were significantly older than those without M-CHIP mutations (60 vs 50 years old). This difference may relate to differences in disease biology between young and older patients with hematologic malignancies, where those who are diagnosed young have a non-aging associated cancer but some other pathogenic mechanism (e.g., inherited predisposition^52^). Clinical implications for higher rates of CH in tumors include potential for accelerated rates of cancer progression, impact on treatment-related outcomes, increased risk for adverse events including secondary cancers, and potential for inferior survival.^53–55^ These differences may be important to consider for precision treatment or management selection for affected patients.

A notable strength of the gnomAD database used in this study is that it provides more weight on genetic diversity compared to other commonly used datasets. For example, the UK Biobank dataset includes 2,782 and 2,364 individuals of South Asian and East Asian continental ancestry, respectively, compared to 15,263 and 8,846 individuals in gnomAD.^56^ The number of African ancestry individuals is also higher in gnomAD (7,451 vs 6,653 in UK Biobank). The analysis presented also has limitations due to the use of secondary aggregate-level data. For example, SM and CH identification were based on population-level AB and age skewing and VAFs in individuals could not be determined. We are also unable to investigate co-inheritance of variants, which is interesting for understanding germline predisposition to SM and CH across ancestry groups.

In summary, population-based genetic databases commonly contain somatic variant data, which can present noise for germline association studies, but provide a good resource for discovery-based research. We used the gnomAD database to create a catalogue of SM and CH variants observed across diverse ancestry groups, which can be used as a reference for future research (e.g., CH targeted sequencing panel design) and as a starting point for quantifying the impact of SM and CH across ancestry groups. We show differences in the frequency and characteristics of SM and CH across ancestries, which are important to consider in study design and disease biology or risk. For example, it may be important to consider broadening of common CHIP sequencing gene lists, particularly for studies that include individuals from typically underrepresented ancestry groups. We show that SM and CH are common across diverse ancestry groups and frequently detected in tumor samples. It is critical to further elucidate differences and quantify risk across variants and racial and ethnic populations to broadly improve precision prevention and patient management.

## Supporting information

Table S3 List of SM and CH variants

## Data Availability

All data produced in the present work are contained in publicly available databases.

## Acknowledgments

This research was made possible through support from H. Lee Moffitt Cancer Center & Research Institute, an NCI designated Comprehensive Cancer Center (P30-CA076292).

## AUTHOR CONTRIBUTIONS

NG and MT designed the research study and co-supervised the project. CCL and YT analyzed the data and created the tables and figures. CCL, NG, and MT wrote the manuscript. All authors reviewed and approved the final manuscript.

## CONFLICTS OF INTEREST

The authors declare no competing financial interests.

## Notes

### Competing Interest Statement

The authors have declared no competing interest.

