## Supplementary Tables and Figures for "Peripheral blood somatic mosaicism and clonal hematopoiesis across ancestry backgrounds"

**SUPPLEMENTARY INFORMATION**

**Table S1. Population in gnomAD exomes (v2) data.**

| Population, n | Exomes | |
|---|---|---|
| | Overall | Non-cancer |
| African/African American | 8,128 | 7,451 |
| Ashkenazi Jewish | 5,040 | 4,786 |
| East Asian | 9,197 | 8,846 |
| European (Finnish) | 10,824 | 10,816 |
| European (non-Finnish) | 56,885 | 51,377 |
| Latino/Admixed American | 17,296 | 17,130 |
| South Asian | 15,308 | 15,263 |
| Other | 3,070 | 2,810 |
| Female (XX) | 57,787 | 53,850 |
| Male (XY) | 67,961 | 64,629 |
| **Total** | **125,748** | **118,479** |

**Table S2. List of regions examined to categorize myeloid and lymphoid clonal hematopoiesis of indeterminate potential** (M-CHIP and L-CHIP, respectively).

| Gene | CH category | Mutations or regions included |
|---|---|---|
| ***ASXL1*** | M-CHIP | Frameshift/nonsense/splice-site in exon 11-12 |
| ***ASXL2*** | M-CHIP | Frameshift/nonsense/splice-site in exon 11-12 |
| ***BCOR*** | M-CHIP | Frameshift/nonsense/splice-site |
| ***BCORL1*** | M-CHIP | Frameshift/nonsense/splice-site |
| ***BRAF*** | M-CHIP | Missense in aa range p. 464, 466, 469, 471, 472, 485, 581, 582, 590-616, 618 |
| ***BRCC3*** | M-CHIP | Frameshift/nonsense/splice-site |
| ***CBL*** | M-CHIP | Missense in Linker/RING finger domains p.345-434 |
| ***CBLB*** | M-CHIP | RING finger missense p.372-412 |
| ***CEBPA*** | M-CHIP | Frameshift/nonsense/splice-site |
| ***CREBBP*** | M-CHIP | Frameshift/nonsense/splice-site, D1435E, R1446L, R1446H, R1446C, Y1450C, P1476R, Y1482H, H1487Y, W1502C, Y1503D, Y1503H, Y1503F, S1680del |
| ***CSF1R*** | M-CHIP | L301F, L301S, Y969C, Y969N, Y969F, Y969H, Y969D |
| ***CSF3R*** | M-CHIP | T615A, T618I, truncating c.741-791 |
| ***CTCF*** | M-CHIP | Frameshift/nonsense/splice-site, R377C, R377H, P378A, P378L |
| ***CUX1*** | M-CHIP | Frameshift/nonsense/splice-site |
| ***DNMT3A*** | M-CHIP | Frameshift/nonsense/splice-site; Missense in aa range: p.290,292-350, 366, 368,379,407,414,462,468,482-614, 634-912 |
| ***EED*** | M-CHIP | Frameshift/nonsense/splice-site, L240Q, I363M |
| ***EP300*** | M-CHIP | Frameshift/nonsense/splice-site, VF1148_1149del, D1399N, D1399Y, P1452L, Y1467N, Y1467H, Y1467C, R1627W, A1629V |
| ***ETNK1*** | M-CHIP | N244S, N244T, N244K |
| ***ETV6*** | M-CHIP | Frameshift/nonsense/splice-site |
| ***EZH2*** | M-CHIP | Frameshift/nonsense/splice-site, Missense in aa range: 62, 102, 145, 159, 164, 202, 238, 244, 283, 292, 488, 497, 561, 568, 617-732, and 740 |
| ***FLT3*** | M-CHIP | V579A, V592A, V592I, F594L, FY590-591GD, D835Y, D835H, D835E, del835 |
| ***GATA1*** | M-CHIP | Frameshift/nonsense/splice-site |
| ***GATA2*** | M-CHIP | Frameshift/nonsense/splice-site, R293Q, N317H, A318T, A318V, A318G, G320D, L321P, L321F, L321V, Q328P, R330Q, R361L, L359V, A372T, R384G, R384K |
| ***GATA3*** | M-CHIP | Frameshift/nonsense/splice-site ZNF domain, R276W, R276Q, N286T, L348V, |
| ***GNA13*** | M-CHIP | I34T, G57S, S62F, M68K, Q134R, Y145F, L152F, E167D, Q169H, R264H, E273K, V322G, V362G, L371F |
| ***GNAS*** | M-CHIP | R201S, R201C, R201H, R201L, Q227K, Q227R, Q227L, Q227H, R374C |
| ***GNB1*** | M-CHIP | G53, K57N, K57M, K57E, K57T, I80T, I80N, I81 |
| ***IDH1*** | M-CHIP | R132C, R132G, R132H, R132L, R132P, R132V, V178I |
| ***IDH2*** | M-CHIP | R140W, R140Q, R140L, R140G, R172W, R172G, R172K, R172T, R172M, R172N, R172S |
| ***IKZF1*** | M-CHIP | Frameshift/nonsense |
| ***IKZF2*** | M-CHIP | Frameshift/nonsense |
| ***IKZF3*** | M-CHIP | Frameshift/nonsense |
| ***JAK1*** | M-CHIP | T478A, T478S, V623A, A634D, L653F, R724H, R724Q, R724P, T782M, L783F |

| | | |
|---|---|---|
| ***JAK2*** | M-CHIP | Missense/indel in aa range: p.536-547, N533D, N533Y, N533S, H538R, K539E, K539L, I540T, I540V, V617F, R683S, R683G, del/ins537-539L, del/ins538-539L, del/ins540-543MK, del/ins540-544MK, del/ins541-543K, del542-543, del543-544, ins11546-547 |
| ***JAK3*** | M-CHIP | M511T, M511I, A572V, A572T, A573V, R657Q, V715I, V715A |
| ***KDM6A*** | M-CHIP | Frameshift/nonsense/splice-site, del419 |
| ***KIT*** | M-CHIP | ins503, V559A, V559D, V559G, V559I, V560D, V560A, V560G, V560E, del560, E561K, del579, P627L, P627T, R634W, K642E, K642Q, V654A, V654E, H697Y, H697D, E761D, K807R, D816H, D816Y, D816F, D816I, D816V, D816H, del551-559 |
| ***KMT2A*** | M-CHIP | Frameshift/nonsense/splice-site |
| ***KMT2D*** | M-CHIP | Frameshift/nonsense |
| ***KRAS*** | M-CHIP | G12D, G12A, G12E, G12V, G13D, G13C, G13Y, G13F, G13R, G13A, G13V, G13E, V14I, T58I, G60D, G60A, G60V, Q61K, Q61E, Q61P, Q61R, Q61L, Q61H, K117E, K117N, A146T, A146P, A146V |
| ***LUC7L2*** | M-CHIP | Frameshift/nonsense/splice-site |
| ***MPL*** | M-CHIP | S505G, S505N, S505C, L510P, del513, W515A, W515R, W515K, W515S, W515L, A519T, A519V, Y591D, W515-518KT |
| ***NF1*** | M-CHIP | Frameshift/nonsense |
| ***NPM1*** | M-CHIP | Frameshift p.W288fs (insertion at c.859_860, 860_861, 862_863, 863_864) |
| ***NRAS*** | M-CHIP | G12S, G12R, G12C, G12N, G12P, G12Y, G12D, G12A, G12V, G12E, G13S, G13R, G13C, G13N, G13P, G13Y, G13D, G13A, G13V, G13E, G60E, G60R, Q61R, Q61L, Q61K, Q61P, Q61H, Q61Q |
| ***PDS5B*** | M-CHIP | Frameshift/nonsense/splice-site, R1292Q |
| ***PDSS2*** | M-CHIP | Frameshift/nonsense |
| ***PHF6*** | M-CHIP | Frameshift/nonsense/splice-site, A40D, M125I, S246Y, F263L, R274Q, C297Y, H302Y, H329L |
| ***PHIP*** | M-CHIP | Frameshift/nonsense/splice-site |
| ***PPM1D*** | M-CHIP | Frameshift/nonsense/splice-site in exon 5/6 |
| ***PRPF40B*** | M-CHIP | Frameshift/nonsense/splice-site, P15H, M58I, P405L, P562S, |
| ***PRPF8*** | M-CHIP | M1307I, C1594W, D1598Y, D1598N, D1598V |
| ***PTEN*** | M-CHIP | Frameshift/nonsense/splice-site, D24G, R47G, F56V, L57W, H61R, K66N, Y68H, C71Y, F81C, Y88C, D92G, D92V, D92E, H93Y, H93D, H93Q, N94I, P95L, I101T, C105F, C105S, D107Y, L112V, H123Y, C124R, C124S, K125E, A126D, K128N, R130G, R130Q, R130L, G132D, I135V, I135K, C136R, C136F, K144Q, A151T, D153Y, D153N, Y155H, Y155C, R159K, R159S, R161K, R161I, G165R, G165E, S170N, S170I, R173C, Y174D, Y177C, H196Y, R234W, G251C, D252Y, F271S, D326G |
| ***PTPN11*** | M-CHIP | Missense in aa range p.58-76, 139, 308, 339, 491-510 |
| ***RAD21*** | M-CHIP | Frameshift/nonsense/splice-site, R65Q, H208R, Q474R |
| ***RUNX1*** | M-CHIP | Frameshift/nonsense/splice-site, S73F, H78Q, H78L, R80C, R80P, R80H, L85Q, P86L, P86H, S114L, D133Y, L134P, R135G, R135K, R135S, R139Q, R142S, A165V, R174Q, R177L, R177Q, A224T, D171G, D171V, D171N, R205W, R223C |
| ***SETBP1*** | M-CHIP | D868N, D868T, S869N, G870S, I871T, D880N, D880Q |
| ***SETD2*** | M-CHIP | Frameshift/nonsense, V1190M |
| ***SETDB1*** | M-CHIP | Frameshift/nonsense, K715E |

| | | |
|---|---|---|
| ***SF1*** | M-CHIP | Frameshift/nonsense/splice-site, T454M, Y476C, A508G |
| ***SF3A1*** | M-CHIP | Frameshift/nonsense/splice-site, A57S, M117I, K166T, Y271C |
| ***SF3B1*** | M-CHIP | Missense in terminal HEAT domains (p.529-1201), and 347, 387 |
| ***SMC1A*** | M-CHIP | Missense at R96, K190T, R586W, M689V, R807H, R1090H, R1090C |
| ***SMC3*** | M-CHIP | Frameshift/nonsense/splice-site, R155I, Q367E, D392V, K571R, R661P, G662C |
| ***SRSF2*** | M-CHIP | Y44H, P95H, P95L, P95T, P95R, P95A, P107H, P95fs |
| ***STAG1*** | M-CHIP | Frameshift/nonsense/splice-site, H1085Y |
| ***STAG2*** | M-CHIP | Frameshift/nonsense/splice-site |
| ***SUZ12*** | M-CHIP | Frameshift/nonsense |
| ***TET2*** | M-CHIP | Frameshift/nonsense/splice-site, missense mutations in catalytic domains (p.1104-1481 and 1843-2002) |
| ***TP53*** | M-CHIP | Frameshift/nonsense/splice-site; Missense in aa range: p.46, 72, 95-288, 320, 330, 334, 347, 348, and 337 |
| ***U2AF1*** | M-CHIP | D14G, S34F, S34Y, R35L, R156H, R156Q, Q157R, Q157P |
| ***U2AF2*** | M-CHIP | Missense in aa range: p. 18, 143, 144, 149-231, p.259-337, p.381-462 |
| ***WT1*** | M-CHIP | Frameshift/nonsense/splice-site |
| ***ZRSR2*** | M-CHIP | Frameshift/nonsense/splice-site, R126P, E133G, C181F, H191Y, I202N, F239V, F239Y, N261Y, C280R, C302R, C326R, H330R, N382K |
| ***CALR*** | M-CHIP | Frameshift in exon 9 |
| ***MYD88*** | M-CHIP | L265P |
| ***NOTCH1*** | M-CHIP | Frameshift/nonsense/splice-site/missense in exon 26-34 |
| ***PIGA*** | M-CHIP | Frameshift/nonsense/splice-site |
| ***STAT3*** | M-CHIP | Missense in SH2 domain (p.580-670) |
| ***ZBTB33*** | M-CHIP | Missense in aa range: p. 22-114,508-531,538-560 |
| ***ACTG1*** | L-CHIP | Splice site |
| ***ARID1A*** | L-CHIP | Frameshift, nonsense, splice site, AAS G2087R |
| ***ARID1B*** | L-CHIP | Frameshift, nonsense, splice site |
| ***ARID5B*** | L-CHIP | Frameshift, nonsense, splice site |
| ***ATM*** | L-CHIP | Frameshift, nonsense, splice site, AAS R337H, R2832H/C, N2875S, I2888T, R3008C/H |
| ***ATR*** | L-CHIP | Frameshift, nonsense, splice site |
| ***B2M*** | L-CHIP | Frameshift, nonsense, splice site, AAS L7V L12P/Q/R |
| ***BCL10*** | L-CHIP | Frameshift, nonsense, splice site |
| ***BCL2*** | L-CHIP | AAS K22N/R, G33R/E/A, P95A/S/L/R, G101V, A131V/D/G/T, N172D/S |
| ***BCL6*** | L-CHIP | AAS R618C |
| ***BIRC3*** | L-CHIP | Frameshift, nonsense, splice site |
| ***BTG1*** | L-CHIP | Frameshift, nonsense, splice site |
| ***BTK*** | L-CHIP | AAS T316A |
| ***CARD11*** | L-CHIP | AAS C49Y/S, E93D/Q, R113Q, F115I/L/V, T117P/N, g123C/D/S, G126D, F130C/I/V, K215E/M/N/T, D230N, S250P, L251P, R337Q, D357E/V, Y361H, D387A, D401N/V, R423W, E626K |
| ***CCND1*** | L-CHIP | AAS S41T, V42E, Y44C/D/F/H/N/S, K46E/I/N, C47R/S, T286A, P287S |
| ***CCND3*** | L-CHIP | Frameshift, nonsense, splice site at C-terminal end, AAS T283A/I, P284S/R/A/L/T, D286G, I290K/R/T/M |

| | | |
|---|---|---|
| ***CD58*** | L-CHIP | Frameshift, nonsense, splice site |
| ***CD79B*** | L-CHIP | AAS Y196H/S/C/N/D/F |
| ***CDKN2A*** | L-CHIP | Frameshift, nonsense, splice site, AAS R112H |
| ***CHEK2*** | L-CHIP | Frameshift, nonsense, splice site |
| ***CIITA*** | L-CHIP | Frameshift, nonsense, splice site, T636M |
| ***CXCR4*** | L-CHIP | Frameshift, nonsense, splice site at C-terminal end |
| ***DDX3X*** | L-CHIP | Frameshift, nonsense, splice site, AAS Y525H, R528H, R534H/C |
| ***DTX1*** | L-CHIP | Frameshift, nonsense, splice site |
| ***ERBB4*** | L-CHIP | AAS K1223T |
| ***FAS*** | L-CHIP | Frameshift, nonsense, splice site |
| ***FAT1*** | L-CHIP | Frameshift, nonsense, splice site, AAS R1627Q, A4419V |
| ***FBXW7*** | L-CHIP | Frameshift, nonsense, splice site, AAS R465H/C/L, Y545C |
| ***FOXO1*** | L-CHIP | Frameshift, nonsense, splice site, AAS S22P/W, T24I/A, S205N |
| ***GPS2*** | L-CHIP | Frameshift, nonsense, splice site |
| ***HIST1H1B*** | L-CHIP | Frameshift, nonsense, splice site |
| ***HIST1H1C*** | L-CHIP | AAS A180P |
| ***HIST1H2BC*** | L-CHIP | AAS E77G/K, A78P, L103F/I |
| ***ID3*** | L-CHIP | Frameshift, nonsense, splice site, AAS P56S/L/R, L64F/H/R/V |
| ***IRF4*** | L-CHIP | AAS C99R |
| ***IRF8*** | L-CHIP | Frameshift, nonsense, splice site at C-terminal end, AAS T80A |
| ***JAK1*** | L-CHIP | AAS Y652H, L910P, Y1035C |
| ***JAK3*** | L-CHIP | AAS 573V, R657Q |
| ***KLF2*** | L-CHIP | Frameshift, nonsense, splice site |
| ***KMT2C*** | L-CHIP | Frameshift, nonsense, splice site |
| ***KMT2D*** | L-CHIP | Frameshift, nonsense, splice site, AAS R5179C, R5432W/Q |
| ***MEF2B*** | L-CHIP | AAS K4E, Y69N, E77K, N81K/Y, D83V |
| ***MGA*** | L-CHIP | Frameshift, nonsense, splice site |
| ***MTOR*** | L-CHIP | AAS W1456G, A1459V, C1483Y, A1971T, T1977K, T1977R, V2006I, S2215F, R2217W, S2231L, I2500F |
| ***MYC*** | L-CHIP | AAS V7L/A/M, T73I/N/A, P74S/A, S161L |
| ***MYD88*** | L-CHIP | AAS M232T, S243N, L265P |
| ***NCOR1*** | L-CHIP | Frameshift, nonsense, splice site |
| ***NFKBIA*** | L-CHIP | Frameshift, nonsense, splice site |
| ***NOTCH1*** | L-CHIP | Frameshift, nonsense, splice site at C-terminal end |
| ***NOTCH2*** | L-CHIP | Frameshift, nonsense, splice site at C-terminal end |
| ***NSD2*** | L-CHIP | AAS E1099K, T1150A |
| ***PAX5*** | L-CHIP | Frameshift, nonsense, splice site, AAS V26G |
| ***PIK3CA*** | L-CHIP | AAS R108H |
| ***PIK3CD*** | L-CHIP | AAS G124D, C416R, E525K, E1021K |
| ***PIM1*** | L-CHIP | AAS L2F, N7K, K24N, G28D/A/S/V, Q37H, H68Y, E79D/K, P81S/A/I/T, S97N/T, L193F/I |
| ***POT1*** | L-CHIP | Frameshift, nonsense, splice site |

| | | |
|---|---|---|
| ***PRDM1*** | L-CHIP | Frameshift, nonsense, splice site |
| ***PTEN*** | L-CHIP | Frameshift, nonsense, splice site, AAS S10N, L23S, D24E, Y27N, L42F, V45I, H123Y, G129R, I135T, Y155S, R335Q |
| ***PTPRD*** | L-CHIP | Frameshift, nonsense, splice site, AAS P1311S, V1565I, R1674H |
| ***RB1*** | L-CHIP | Frameshift, nonsense, splice site, AAS R661W, C706R, S758L |
| ***RHOA*** | L-CHIP | AAS R5Q/W, G17A, Y34N, N41S, Y42C/F/S/H, Y66N, L69P/R, A161T |
| ***SMARCA4*** | L-CHIP | Frameshift, nonsense, splice site, AAS T910M/A, R973Q/L, R1135Q, R1189L, R1192C/H, R1243L/W |
| ***SOCS1*** | L-CHIP | Frameshift, nonsense, splice site, AAS A3T/S/P/V |
| ***SPEN*** | L-CHIP | Frameshift, nonsense, splice site |
| ***STAT3*** | L-CHIP | AAS in SH2 domain (S614N/R, E616G/K, Y640F, K658R, D661V/Y) |
| ***TBL1XR1*** | L-CHIP | AAS D370N, W376C, Y395C/H |
| ***TNFAIP3*** | L-CHIP | Frameshift, nonsense, splice site |
| ***TNFRSF14*** | L-CHIP | Frameshift, nonsense, splice site |
| ***TSC2*** | L-CHIP | Frameshift, nonsense, splice site |
| ***XPO1*** | L-CHIP | AAS E571K/V/Q/G |

**Table S3. List of gnomAD identified somatic mosaicism (SM) and clonal hematopoiesis (CH) variants.** All SM variants identified by our pipeline are listed (column A) and those categorized as CH are indicated (column B). Variants in known pathogenic myeloid (M-CHIP) and lymphoid (L-CHIP) regions are noted (column C and D, respectively). Functional characterizations for each variant are presented (columns E-H), and those present in all ancestry populations are noted (column I).

See Excel file “Table S3 List of SM and CH variants”

**Table S4. Somatic mosaicism (SM) and clonal hematopoiesis (CH) variant characteristics.**

| | SM<br>N = 503,703 | CH<br>N = 89,361 |
|---|---|---|
| **Variant effect predictor annotation (gnomAD)** | | |
| **Somatic, n (%)** | | |
| No | 472,362 (94%) | 84,358 (94%) |
| Yes | 31,341 (6.2%) | 5,003 (5.6%) |
| **CADD PHRED categories, n (%)** | | |
| ≥ 20, Likely pathogenic | 137,286 (64%) | 24,751 (65%) |
| < 20 | 77,722 (36%) | 13,232(35%) |
| Unknown | 288,695 | 51,378 |
| **dbGap PopFreq, median (IQR)** | | |
| Reference allele | 1 (0.99, 1.0) | 1 (0.99, 1.0) |
| Unknown | 124,495 | 20,140 |
| Alternate allele | 0 (0, 0.0001) | 0 (0, 0.0001) |
| Unknown | 166,239 | 27,604 |
| **Cumulative variant allele count, n** | | |
| **M-CHIP** | | |
| *DNMT3A* | 365 | 214 |
| *CUX1* | 122 | 0 |
| *SF3B1* | 114 | 46 |
| *NF1* | 103 | 0 |
| *TP53* | 98 | 7 |
| *JAK2* | 88 | 88 |
| *PDS5B* | 58 | 42 |
| *MFSD11* | 50 | 50 |
| *EZH2* | 50 | 0 |
| *ASXL1* | 41 | 25 |
| *GNB1* | 25 | 25 |
| *KMT2D* | 24 | 10 |
| *WT1* | 23 | 0 |
| *TET2* | 17 | 0 |
| *PRPF40B* | 12 | 12 |
| **L-CHIP** | | |
| *TSC2* | 32 | 3 |
| *SMARCA4* | 24 | 23 |
| *KMT2D* | 24 | 10 |
| *PTPRD* | 15 | 2 |
| *ARID5B* | 15 | 0 |
| *NOTCH1* | 11 | 6 |

IQR, interquartile range.

Genes in M-CHIP and L-CHIP with more than 10 variants are included

**Table S5. Genes with the highest prevalence (cumulative variant allele count) of somatic mosaicism (SM) and clonal hematopoiesis (CH) variants.**

| SM | | CH | |
|---|---|---|---|
| **Gene** | **Cumulative variant allele count, n** | **Gene** | **Cumulative variant allele count, n** |
| *MUC3A* | 6,894,968 | *ARID3A* | 4,104 |
| *HLA-C* | 5,705,906 | *FAT3* | 2,975 |
| *MUC6* | 3,706,059 | *OPLAH* | 2,660 |
| *HLA-DQA1* | 2,927,734 | *ECSIT* | 2,332 |
| *OR9G1* | 2,793,939 | *GAL3ST1* | 2,237 |
| *MAP2K3* | 2,515,390 | *LMF1* | 1,565 |
| *HLA-B* | 2,466,511 | *TSHZ3* | 1,515 |
| *OR8U1* | 2,124,261 | *GGT5* | 1,493 |
| *HLA-DQB1* | 2,121,434 | *SLC22A2* | 1,208 |
| *PSMD13* | 1,721,884 | *SYNJ2* | 1,201 |
| *HLA-DPA1* | 1,499,049 | *SLC16A8* | 1,168 |
| *CYP4F22* | 1,477,031 | *PPFIBP2* | 1,130 |
| *PKD1L2* | 1,033,141 | *PLIN4* | 1,117 |
| *ACSF3* | 991,475 | *ADAMTSL1* | 1,066 |
| *HLA-DPB1* | 950,617 | *RNF4* | 1,024 |
| *OR4L1* | 844,406 | *MUC16* | 1,016 |
| *SIRPA* | 806,597 | *HNF1B* | 984 |
| *MPHOSPH6* | 691,800 | *OR51C1P* | 855 |
| *PCDHA4* | 679,079 | *GLE1* | 688 |
| *CDC27* | 678,397 | *OBSCN* | 675 |

**Table S6. Scaled population allele frequencies (sPAF) of somatic mosaicism (SM) and clonal hematopoiesis (CH) variants in the study population**. Data are presented by ancestry groups and sex. Medians are calculated using the sPAF of all variants with non-zero values for each population.

| | SM | | CH | |
|---|---|---|---|---|
| | **sPAF, median (IQR)** | **p-value** | **sPAF, median (IQR)** | **p-value** |
| **All gnomAD populations** | 2.69e-04 (1.35e-04, 5.29e-04) | | 2.03e-04 (1.04e-04, 3.64e-04) | |
| **Ancestry** | | | | |
| African/African American | 1.51e-04 (7.95e-05, 4.86e-04) | **<0.001** | 1.02e-04 (7.39e-05, 1.81e-04) | **<0.001** |
| Ashkenazi Jewish | 1.97e-04 (7.27e-05, 9.36e-04) | | 1.31e-04 (7.04e-05, 3.94e-04) | |
| East Asian | 1.35e-04 (7.37e-05, 3.96e-04) | | 9.61e-05 (7.17e-05, 1.98e-04) | |
| European (Finnish) | 1.97e-04 (8.02e-05, 8.55e-04) | | 1.06e-04 (7.3e-05, 2.13e-04) | |
| European (non-Finnish) | 1.55e-04 (8.43e-05, 3.33e-04) | | 1.39e-04 (8.06e-05, 2.61e-04) | |
| Latino/Admixed American | 1.41e-04 (7.54e-05, 3.59e-04) | | 1.11e-04 (7.25e-05, 2.06e-04) | |
| South Asian | 1.33e-04 (7.51e-05, 3.32e-04) | | 8.4e-05 (7.11e-05, 1.44e-04) | |
| Other | 8.73e-05 (6.79e-05, 2.62e-04) | | 7.39e-05 (6.7e-05, 9.42e-05) | |
| **Sex** | | | | |
| Female | 1.71e-04 (8.8e-05, 3.35e-04) | **<0.001** | 1.38e-04 (8.09e-05, 2.31e-04) | 0.2 |
| Male | 1.95e-04 (9.23e-05, 3.57e-04) | | 1.37e-04 (8.09e-05, 2.31e-04) | |

**Table S7. Scaled population allele frequencies (sPAF) of somatic mosaicism (SM) and clonal hematopoiesis (CH) variants in curated gene regions**. Data are presented based on in curated myeloid (M-CHIP) and lymphoid (L-CHIP) gene regions. Medians are calculated using the sPAF of all variants with non-zero values for each population.

| | SM | | | | CH | | | |
|---|---|---|---|---|---|---|---|---|
| | M-CHIP | | L-CHIP | | M-CHIP | | L-CHIP | |
| | **sPAF, median (IQR)** | **p-value** | **sPAF, median (IQR)** | **p-value** | **sPAF, median (IQR)** | **p-value** | **sPAF, median (IQR)** | **p-value** |
| **All gnomAD populations** | 2.62e-04 (1.31e-04, 5.24e-04) | | 1.15e-04 (6.97e-05, 3.15e-04) | | 2.62e-04 (1.31e-04, 4.59e-04) | | 9.18e-05 (6.97e-05, 3.31e-04) | |
| **Ancestry** | | | | | | | | |
| African/African American | 1.18e-04 (6.56e-05, 2.63e-04) | | 2.14e-04 (7.25e-05, 2.63e-04) | | 6.68e-05 (6.56e-05, 1.97e-04) | | 7.62e-05 (6.63e-05, 2.63e-04) | |
| Ashkenazi Jewish | 6.67e-05 (6.56e-05, 1.97e-04) | | 1.59e-04 (1.1e-04, 2.08e-04) | | 6.57e-05 (6.56e-05, 1.97e-04) | | 1.1e-04 (1.1e-04, 1.1e-04) | |
| East Asian | 6.79e-05 (6.55e-05, 1.97e-04) | | 8.55e-05 (7.49e-05, 1.42e-04) | | 6.56e-05 (6.55e-05, 1.31e-04) | | 6.55e-05 (6.55e-05, 6.55e-05) | |
| European (Finnish) | 6.85e-05 (6.56e-05, 2.26e-04) | **<0.001** | 6.85e-05 (6.59e-05, 8.36e-05) | 0.13 | 6.66e-05 (6.56e-05, 1.48e-04) | **<0.001** | 6.7e-05 (6.59e-05, 6.85e-05) | 0.4 |
| European (non-Finnish) | 1.97e-04 (7.58e-05, 3.94e-04) | | 8.35e-05 (6.71e-05, 2.46e-04) | | 1.97e-04 (1.06e-04, 3.31e-04) | | 1.68e-04 (6.67e-05, 3.7e-04) | |
| Latino/Admixed American | 6.62e-05 (6.56e-05, 1.51e-04) | | 1.15e-04 (8.3e-05, 3.52e-04) | | 6.57e-05 (6.56e-05, 1.31e-04) | | 8.3e-05 (6.67e-05, 9.1e-05) | |
| South Asian | 8.77e-05 (6.56e-05, 1.48e-04) | | 1.09e-04 (7.45e-05, 2.03e-04) | | 6.56e-05 (6.55e-05, 1.32e-04) | | 1.45e-04 (8.77e-05, 2.03e-04) | |
| Other | 6.72e-05 (6.56e-05, 1.31e-04) | | 6.65e-05 (6.56e-05, 6.73e-05) | | 6.67e-05 (6.56e-05, 7.33e-05) | | 6.73e-05 (6.73e-05, 6.73e-05) | |
| **Sex** | | | | | | | | |
| Female | 1.38e-04 (7.01e-05, 2.88e-04) | 0.6 | 1.15e-04 (6.95e-05, 3.1e-04) | 0.8 | 1.32e-04 (6.57e-05, 3.02e-04) | 0.4 | 1.35e-04 (7.01e-05, 4e-04) | 0.5 |
| Male | 1.97e-04 (6.78e-05, 3.96e-04) | | 1.32e-04 (6.85e-05, 2.99e-04) | | 1.97e-04 (6.78e-05, 3.65e-04) | | 1.31e-04 (8.66e-05, 1.45e-04) | |

**Table S8. Prevalence of somatic mosaicism (SM) and clonal hematopoiesis (CH) variants within the curated myeloid (M-CHIP) gene lists.** Prevalence is estimated based on the cumulative variant allele count within each gene. AFR, African/African American; ASJ, Ashkenazi Jewish; EAS, East Asian; FIN, European (Finnish); NFE, Non-Finnish European; AMR, Latino/Admixed American; SAS, South Asian; OTH, Other

| | SM in M-CHIP genes (n) | | | | | | | | | M-CHIP (n) | | | | | | | | |
|---|---|---|---|---|---|---|---|---|---|---|---|---|---|---|---|---|---|---|
| **Symbol** | **AFR** | **ASJ** | **EAS** | **FIN** | **NFE** | **AMR** | **SAS** | **OTH** | **All** | **AFR** | **ASJ** | **EAS** | **FIN** | **NFE** | **AMR** | **SAS** | **OTH** | **All** |
| *DNMT3A* | 39 | 13 | 24 | 71 | 161 | 22 | 23 | 12 | 365 | 26 | 11 | 21 | 18 | 101 | 8 | 21 | 8 | 214 |
| *CUX1* | 3 | 5 | 4 | 11 | 45 | 21 | 32 | 1 | 122 | 0 | 0 | 0 | 0 | 0 | 0 | 0 | 0 | 0 |
| *SF3B1* | 29 | 5 | 2 | 16 | 44 | 7 | 7 | 4 | 114 | 1 | 3 | 1 | 2 | 29 | 6 | 3 | 1 | 46 |
| *NF1* | 41 | 2 | 17 | 5 | 34 | 4 | 0 | 0 | 103 | 0 | 0 | 0 | 0 | 0 | 0 | 0 | 0 | 0 |
| *TP53* | 11 | 5 | 4 | 9 | 58 | 3 | 7 | 1 | 98 | 0 | 1 | 0 | 0 | 6 | 0 | 0 | 0 | 7 |
| *JAK2* | 5 | 6 | 3 | 12 | 45 | 6 | 10 | 1 | 88 | 5 | 6 | 3 | 12 | 45 | 6 | 10 | 1 | 88 |
| *PDS5B* | 5 | 3 | 0 | 1 | 44 | 4 | 1 | 0 | 58 | 4 | 3 | 0 | 1 | 30 | 3 | 1 | 0 | 42 |
| *MFSD11* | 5 | 2 | 2 | 0 | 37 | 1 | 2 | 1 | 50 | 5 | 2 | 2 | 0 | 37 | 1 | 2 | 1 | 50 |
| *EZH2* | 9 | 0 | 11 | 7 | 22 | 1 | 0 | 0 | 50 | 0 | 0 | 0 | 0 | 0 | 0 | 0 | 0 | 0 |
| *ASXL1* | 2 | 2 | 5 | 4 | 26 | 1 | 0 | 1 | 41 | 2 | 2 | 2 | 2 | 16 | 1 | 0 | 0 | 25 |
| *GNB1* | 0 | 1 | 2 | 3 | 11 | 5 | 2 | 1 | 25 | 0 | 1 | 2 | 3 | 11 | 5 | 2 | 1 | 25 |
| *KMT2D* | 3 | 0 | 0 | 0 | 11 | 8 | 1 | 1 | 24 | 2 | 0 | 0 | 1 | 6 | 1 | 0 | 0 | 10 |
| *WT1* | 0 | 0 | 0 | 0 | 0 | 0 | 22 | 0 | 23 | 0 | 0 | 0 | 0 | 0 | 0 | 0 | 0 | 0 |
| *TET2* | 0 | 0 | 0 | 1 | 4 | 2 | 11 | 0 | 17 | 0 | 0 | 0 | 0 | 0 | 0 | 0 | 0 | 0 |
| *PRPF40B* | 0 | 3 | 0 | 2 | 2 | 2 | 2 | 1 | 12 | 0 | 3 | 0 | 2 | 2 | 2 | 2 | 1 | 12 |
| *NOTCH1* | 0 | 0 | 0 | 3 | 6 | 0 | 0 | 0 | 9 | 0 | 0 | 0 | 1 | 4 | 0 | 0 | 0 | 5 |
| *PRPF8* | 0 | 0 | 1 | 0 | 6 | 1 | 0 | 0 | 8 | 0 | 0 | 1 | 0 | 6 | 1 | 0 | 0 | 8 |
| *IDH2* | 1 | 0 | 1 | 0 | 6 | 0 | 0 | 0 | 8 | 1 | 0 | 1 | 0 | 6 | 0 | 0 | 0 | 8 |
| *GNAS* | 0 | 1 | 0 | 0 | 2 | 1 | 0 | 0 | 4 | 0 | 1 | 0 | 0 | 2 | 1 | 0 | 0 | 4 |
| *CSDE1* | 0 | 0 | 0 | 1 | 1 | 0 | 0 | 0 | 2 | 0 | 0 | 0 | 1 | 1 | 0 | 0 | 0 | 2 |
| *KMT2A* | 2 | 0 | 0 | 0 | 0 | 0 | 0 | 0 | 2 | 2 | 0 | 0 | 0 | 0 | 0 | 0 | 0 | 2 |
| *SETD2* | 0 | 0 | 0 | 0 | 1 | 0 | 0 | 0 | 1 | 0 | 0 | 0 | 0 | 1 | 0 | 0 | 0 | 1 |
| *CREBBP* | 0 | 0 | 0 | 0 | 0 | 0 | 0 | 0 | 1 | 0 | 0 | 0 | 1 | 0 | 0 | 0 | 0 | 1 |
| *SMC3* | 0 | 0 | 0 | 1 | 1 | 0 | 0 | 0 | 1 | 0 | 0 | 0 | 0 | 0 | 0 | 0 | 0 | 0 |
| *STAT3* | 0 | 0 | 0 | 0 | 1 | 0 | 0 | 0 | 1 | 0 | 0 | 0 | 0 | 0 | 0 | 0 | 0 | 0 |
| *CBL* | 0 | 0 | 0 | 0 | 0 | 1 | 0 | 0 | 1 | 0 | 0 | 0 | 0 | 0 | 0 | 0 | 0 | 0 |
| *RUNX1* | 0 | 0 | 0 | 0 | 0 | 0 | 1 | 0 | 1 | 0 | 0 | 0 | 0 | 0 | 0 | 0 | 0 | 0 |
| **TOTAL** | **155** | **48** | **76** | **147** | **568** | **90** | **121** | **24** | **1229** | **48** | **33** | **33** | **44** | **303** | **35** | **41** | **13** | **550** |

**Table S9. Prevalence of somatic mosaicism (SM) and clonal hematopoiesis (CH) variants within the curated lymphoid (L-CHIP) gene lists.** Prevalence is estimated based on the cumulative variant allele count within each gene. AFR, African/African American; ASJ, Ashkenazi Jewish; EAS, East Asian; FIN, European (Finnish); NFE, Non-Finnish European; AMR, Latino/Admixed American; SAS, South Asian; OTH, Other

| | SM in L-CHIP genes (n) | | | | | | | | | L-CHIP (n) | | | | | | | | |
|---|---|---|---|---|---|---|---|---|---|---|---|---|---|---|---|---|---|---|
| **Symbol** | **AFR** | **ASJ** | **EAS** | **FIN** | **NFE** | **AMR** | **SAS** | **OTH** | **All** | **AFR** | **ASJ** | **EAS** | **FIN** | **NFE** | **AMR** | **SAS** | **OTH** | **All** |
| *TSC2* | 21 | 0 | 0 | 1 | 1 | 8 | 0 | 1 | 32 | 0 | 0 | 0 | 1 | 1 | 0 | 0 | 1 | 3 |
| *SMARCA4* | 4 | 1 | 3 | 1 | 15 | 0 | 0 | 0 | 24 | 4 | 1 | 3 | 1 | 14 | 0 | 0 | 0 | 23 |
| *ARID5B* | 4 | 0 | 3 | 2 | 6 | 0 | 0 | 0 | 15 | 0 | 0 | 0 | 0 | 0 | 0 | 0 | 0 | 0 |
| *KMT2D* | 3 | 0 | 0 | 1 | 11 | 8 | 1 | 0 | 24 | 2 | 0 | 0 | 1 | 6 | 1 | 0 | 0 | 10 |
| *PTPRD* | 4 | 0 | 0 | 0 | 6 | 5 | 0 | 0 | 15 | 0 | 0 | 0 | 0 | 2 | 0 | 0 | 0 | 2 |
| *NOTCH1* | 0 | 0 | 0 | 3 | 6 | 1 | 1 | 0 | 11 | 0 | 0 | 0 | 1 | 4 | 0 | 1 | 0 | 6 |
| *ARID1A* | 0 | 0 | 0 | 0 | 3 | 3 | 3 | 0 | 9 | 0 | 0 | 0 | 0 | 0 | 0 | 0 | 0 | 0 |
| *CHEK2* | 0 | 0 | 0 | 0 | 4 | 1 | 2 | 1 | 8 | 0 | 0 | 0 | 0 | 0 | 0 | 0 | 0 | 0 |
| *KMT2C* | 0 | 0 | 1 | 1 | 4 | 0 | 0 | 1 | 7 | 0 | 0 | 0 | 0 | 0 | 0 | 0 | 0 | 0 |
| *POT1* | 1 | 0 | 0 | 0 | 5 | 0 | 0 | 0 | 6 | 1 | 0 | 0 | 0 | 5 | 0 | 0 | 0 | 6 |
| *ARID1B* | 0 | 1 | 0 | 0 | 2 | 1 | 0 | 0 | 4 | 0 | 1 | 0 | 0 | 0 | 0 | 0 | 0 | 1 |
| *DTX1* | 0 | 0 | 0 | 0 | 2 | 0 | 1 | 0 | 3 | 0 | 0 | 0 | 0 | 1 | 0 | 1 | 0 | 2 |
| *KLF2* | 0 | 1 | 0 | 0 | 0 | 0 | 2 | 0 | 3 | 0 | 0 | 0 | 0 | 0 | 0 | 0 | 0 | 0 |
| *CDKN2A* | 0 | 0 | 0 | 0 | 3 | 2 | 0 | 0 | 5 | 0 | 0 | 0 | 0 | 0 | 0 | 0 | 0 | 0 |
| *PAX5* | 0 | 0 | 0 | 0 | 1 | 1 | 1 | 0 | 3 | 0 | 0 | 0 | 0 | 0 | 1 | 0 | 0 | 1 |
| *SPEN* | 0 | 0 | 0 | 0 | 2 | 0 | 0 | 0 | 2 | 0 | 0 | 0 | 0 | 0 | 0 | 0 | 0 | 0 |
| *BTG1* | 0 | 0 | 1 | 0 | 0 | 0 | 1 | 0 | 2 | 0 | 0 | 0 | 0 | 0 | 0 | 0 | 0 | 0 |
| *MGA* | 0 | 0 | 0 | 0 | 2 | 0 | 0 | 0 | 2 | 0 | 0 | 0 | 0 | 0 | 0 | 0 | 0 | 0 |
| *CIITA* | 0 | 0 | 0 | 1 | 1 | 0 | 0 | 0 | 2 | 0 | 0 | 0 | 1 | 1 | 0 | 0 | 0 | 2 |
| *TNFRSF14* | 2 | 0 | 0 | 0 | 2 | 0 | 1 | 0 | 5 | 0 | 0 | 0 | 0 | 0 | 0 | 0 | 0 | 0 |
| *STAT3* | 0 | 0 | 0 | 0 | 1 | 0 | 0 | 0 | 1 | 0 | 0 | 0 | 0 | 0 | 0 | 0 | 0 | 0 |
| *FBXW7* | 0 | 0 | 0 | 0 | 0 | 0 | 1 | 0 | 1 | 0 | 0 | 0 | 0 | 0 | 0 | 0 | 0 | 0 |
| *CCND3* | 0 | 0 | 0 | 0 | 0 | 1 | 0 | 0 | 1 | 0 | 0 | 0 | 0 | 0 | 1 | 0 | 0 | 1 |
| *PRDM1* | 0 | 0 | 0 | 0 | 0 | 0 | 1 | 0 | 1 | 0 | 0 | 0 | 0 | 0 | 0 | 0 | 0 | 0 |
| *TNFAIP3* | 0 | 0 | 0 | 1 | 0 | 0 | 0 | 0 | 1 | 0 | 0 | 0 | 0 | 0 | 0 | 0 | 0 | 0 |
| *FAT1* | 0 | 0 | 0 | 0 | 0 | 0 | 0 | 0 | 0 | 0 | 0 | 0 | 0 | 0 | 0 | 0 | 0 | 0 |
| **TOTAL** | **39** | **3** | **8** | **11** | **77** | **31** | **15** | **3** | **187** | **7** | **2** | **3** | **5** | **34** | **3** | **2** | **1** | **57** |

**Table S10. Age of patients with and without gnomAD-identified clonal hematopoiesis (CH) variants in tumor sequencing.** CH variants identified in gnomAD were linked with the Catalogue of Somatic Mutations in Cancer (COSMIC) population database. Data for prevalence of CH variants are shown across all cancer types and stratified by hematologic and non-hematologic cancers.

| | **All cancer types** | | | | **Hematologic** | | | | **Non-hematologic** | | | |
|---|---|---|---|---|---|---|---|---|---|---|---|---|
| | **Overall N = 90,191** | **CH variants N = 55,190** | **No CH variants N = 35,001** | **p-value** | **Overall N =53,542** | **CH variants N = 46,744** | **No CH variants N = 6,798** | **p-value** | **Overall N = 36,649** | **CH variants N = 8,446** | **No CH variants N = 28,203** | **p-value** |
| Age, Median (IQR) | 60 (47, 69) | 61 (49, 71) | 59 (47, 69) | **<0.001** | 57 (35, 68) | 60 (45, 70) | 51 (14, 66) | **<0.001** | 60 (49, 70) | 62 (51, 71) | 60 (48, 69) | **<0.001** |
| Age unknown | 62,015 | 46,881 | 15,134 | | 47,752 | 43,558 | 4,194 | | 14,263 | 3,323 | 10,940 | |

Individuals are considered CH carriers if at least one CH mutation is detected.
IQR, interquartile range.

**Table S11. Prevalence of clonal hematopoiesis (CH) detected in tumors.** CH variants identified in gnomAD were linked with the Catalogue of Somatic Mutations in Cancer (COSMIC) population database. COSMIC cancer types with the highest prevalence of CH in tumors are shown.

| | **Overall** | **CH variants** |
|---|---|---|
| Hematopoietic and lymphoid tissue | 53,504 | 46,706 (87%) |
| Large intestine | 4,386 | 1,804 (41%) |
| Endometrium | 877 | 335 (38%) |
| Stomach | 1,497 | 529 (35%) |
| Skin | 2,008 | 633 (32%) |
| Pancreas | 1,903 | 541 (28%) |
| Thyroid | 892 | 236 (26%) |
| Urinary tract | 1,002 | 244 (24%) |
| Esophagus | 1,348 | 322 (24%) |
| Biliary tract | 847 | 185 (22%) |
| Upper aerodigestive tract | 1,516 | 349 (23%) |
| Ovary | 1,161 | 245 (21%) |
| Lung | 3,554 | 604 (17%) |
| Liver | 2,073 | 348 (17%) |
| Central nervous system | 2,366 | 390 (16%) |
| Breast | 3,533 | 581 (16%) |
| Soft tissue | 698 | 83 (12%) |
| Prostate | 2,515 | 295 (12%) |
| Kidney | 1,626 | 150 (9.2%) |

Individuals are considered CH carriers if at least one CH mutation is detected.

Cancer types with more than 500 samples are shown.

**Table S12. Age characteristics of patients with clonal hematopoiesis (CH) in myeloid (M-CHIP) or lymphoid (L-CHIP) regions.** CH variants identified in gnomAD were linked with the Catalogue of Somatic Mutations in Cancer (COSMIC) population database. Data for prevalence of M-CHIP or L-CHIP are shown for all tumor types and stratified by hematologic and non-hematologic cancers included in COSMIC.

| | All tumor types | | | Hematologic | | | Non-hematologic | | |
|---|---|---|---|---|---|---|---|---|---|
| | **Sample size (N)** | **Overall N = 90,191** | **p-value** | **Sample size (N)** | **Overall N = 53,542** | **p-value** | **Sample size (N)** | **Overall N = 36,649** | **p-value** |
| **Age, Median (IQR)** | | | | | | | | | |
| **All CH** | 55,190 | 61 (49, 71) | | 46,762 | 60 (45, 70) | | 8,503 | 62 (51, 71) | |
| **M-CHIP** | | | **<0.001** | | | **<0.001** | | | **<0.001** |
| Yes | 49,905 | 61 (48, 70) | | 46,422 | 60 (46, 70) | | 3,526 | 61 (50, 71) | |
| No | 5,285 | 62 (51, 71) | | 340 | 50 (13, 64) | | 4,977 | 63 (52, 71) | |
| **L-CHIP** | | | 0.4 | | | 0.04 | | | 0.7 |
| Yes | 14 | 50 (37, 68) | | 6 | 19 (11, 28) | | 8 | 66 (55, 72) | |
| No | 55,176 | 61 (49, 71) | | 46,756 | 60 (45, 70) | | 8,495 | 62 (51, 71) | |

Individuals are considered CH carriers if at least one CH mutation is detected.
IQR, interquartile range.

### SUPPLEMENTARY FIGURES

**Figure S1. Circle plot visualizing the decrease in variant counts during successive filtering.**

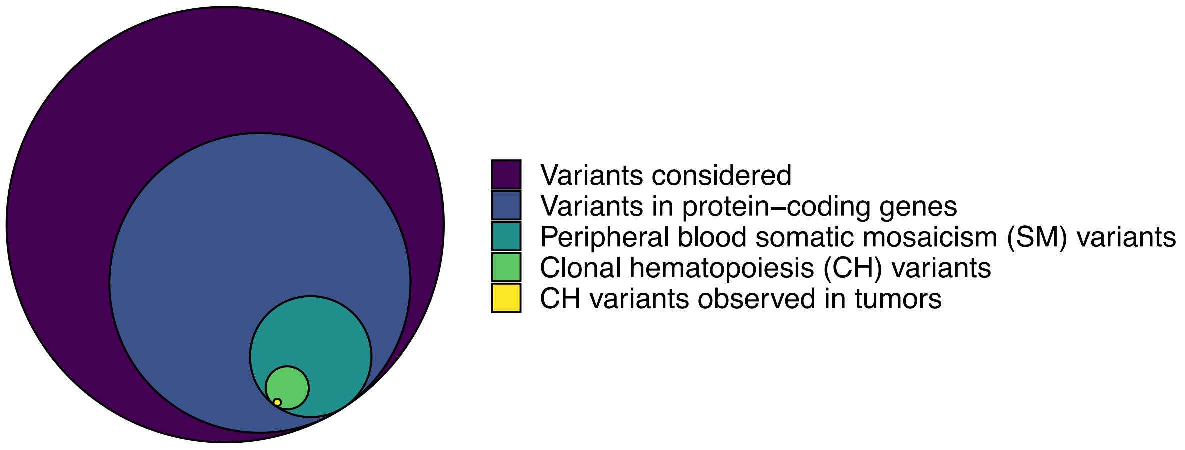

**Figure S2. Mutation types in the gnomAD population overall, somatic mosaicism (SM) variants, and clonal hematopoiesis (CH) variants.** Counts and percentages of mutation types are displayed in the table.

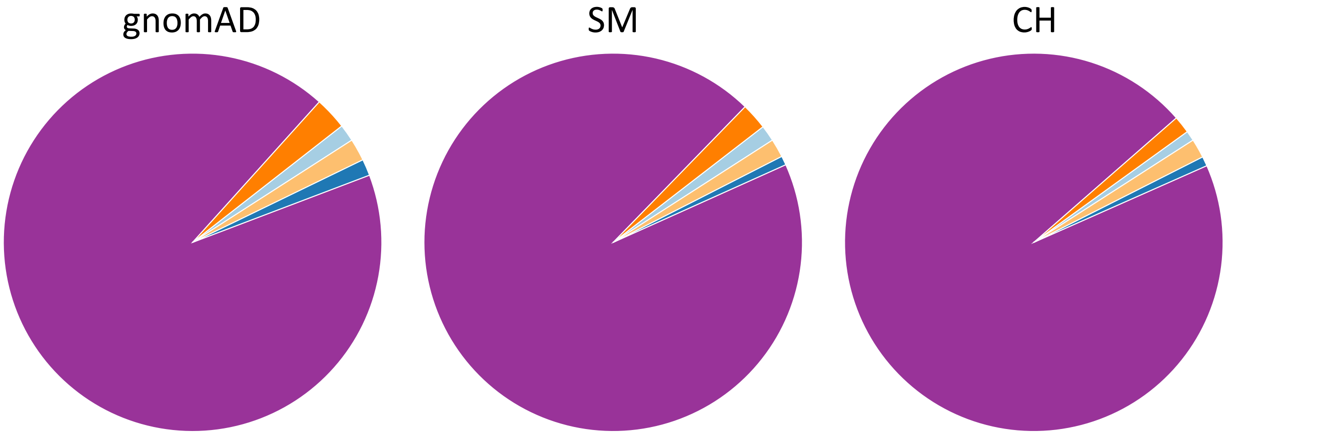


| Nucleotide types | gnomAD | SM variants | CH variants |
|---|---|---|---|
| Large deletion | 57,609 (1.5%) | 6,946 (1.4%) | 772 (0.9%) |
| Large insertion | 109,304 (2.8%) | 11,408 (2.3%) | 1,322 (1.5%) |
| Single nucleotide | 3,637,547 (92.5%) | 473,487 (94%) | 85,083 (95%) |
| Small deletion | 55,538 (1.4%) | 3,917 (0.8%) | 731 (0.8%) |
| Small insertion | 74,543 (1.9%) | 7,945 (1.6%) | 1,453 (1.6%) |

**Figure S3. Scaled population allele frequencies (sPAF) of select myeloid CH (M-CHIP) mutations in the gnomAD population.**

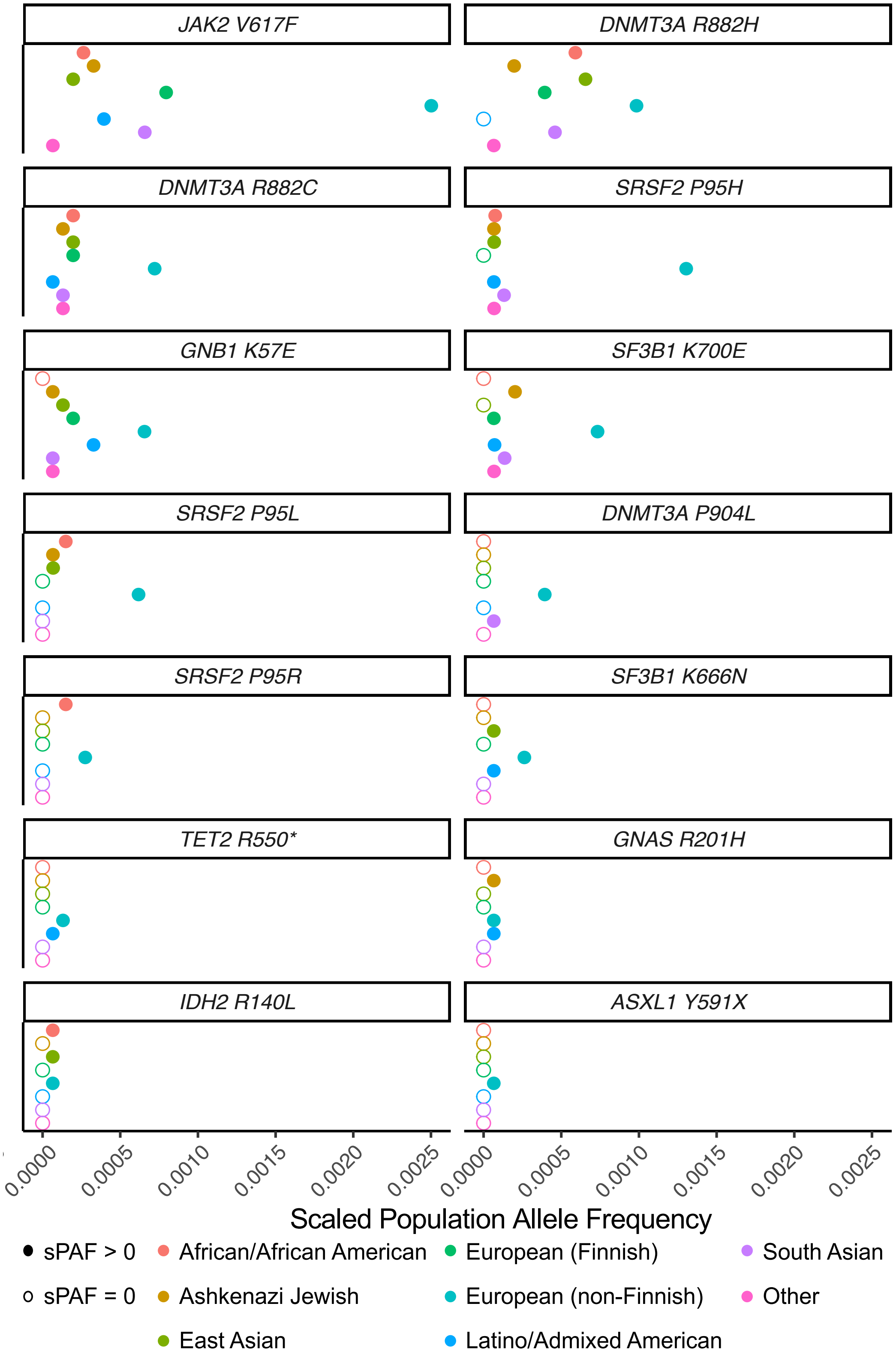

**Figure S4. Age distribution of cancer patients with and without clonal hematopoiesis (CH) variants detected in their tumor sample.** Data is from the COSMIC database. The vertical lines correspond to median ages.

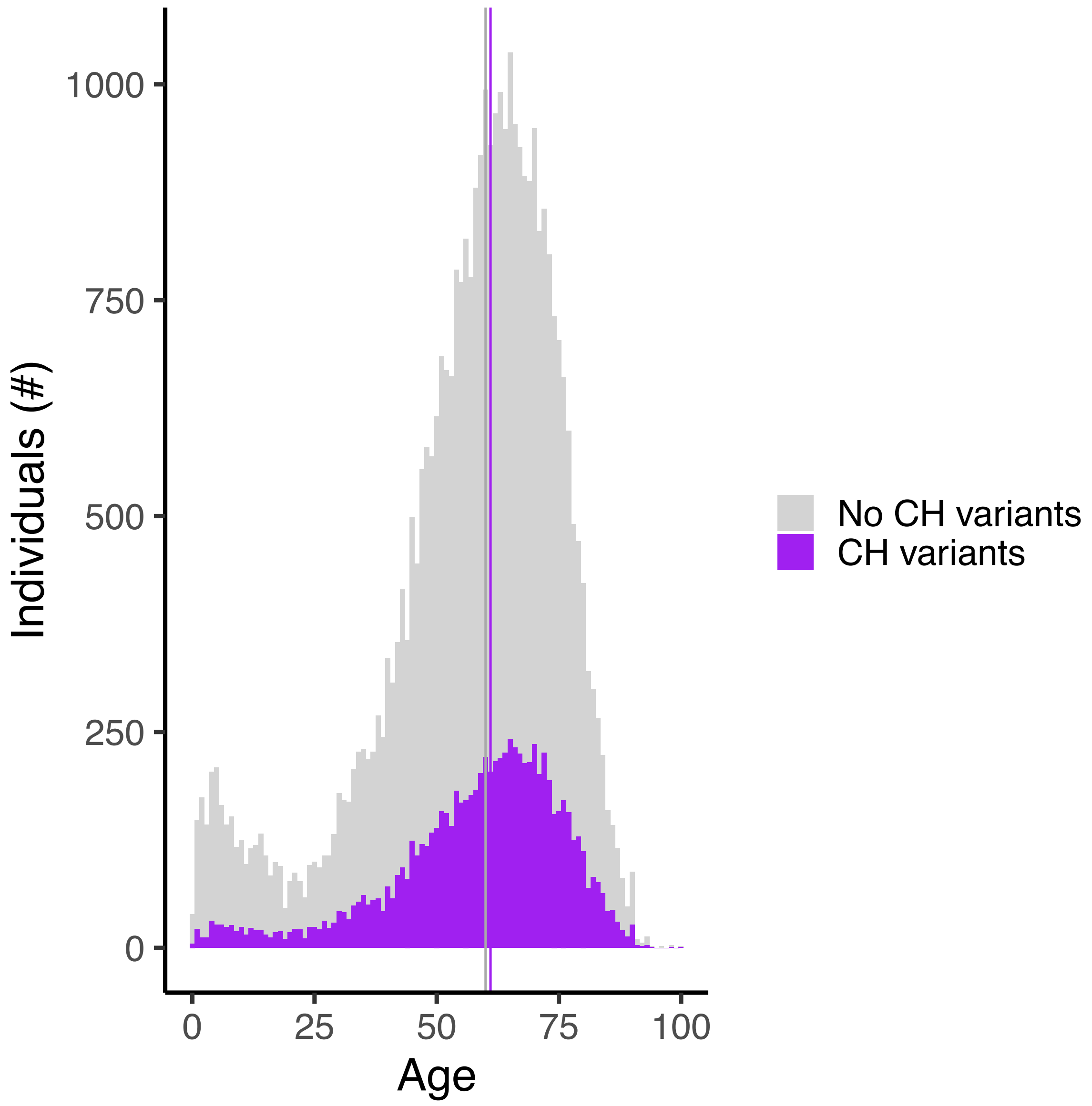
